# Multimodal AI for Precision Preventive Cardiology

**DOI:** 10.1101/2025.10.09.25337677

**Authors:** Devansh Pandey, Liaoyi Xu, Eucharist Kun, Chenfei Li, Joyce Y. Wang, Alaa Melek, Julie C. DiCarlo, Edward Castillo, Jagat Narula, Charles A. Taylor, Vagheesh M. Narasimhan

## Abstract

Coronary artery disease (CAD) is the leading cause of death worldwide, yet it is highly preventable. Early detection is critical, particularly because the first clinical manifestation of CAD is a heart attack in ∼50% of individuals. Current clinical risk scores rely largely on traditional biomarkers and do not leverage recent advances in medical imaging and genetics. To address this, we developed *preCog*, a multimodal artificial intelligence framework that integrates multi-organ imaging, genetic risk, metabolic biomarkers, ECGs and demographic data to predict time to incident CAD over up to 10 years of follow-up in a cohort of 60,000 UK Biobank participants. To quantify imaging risk, we applied 2D and 3D foundational computer vision models to more than 500,000 images spanning cardiac, liver and pancreas MRI, as well as DXA scans, each capturing distinct facets of CAD pathophysiology. We extracted deep learning derived image embeddings and compressed them to compact representations that were highly correlated with conventional imaging metrics of cardiac function (e.g. ejection fraction and stroke volumes) yet outperformed these metrics for CAD prediction (AUC 0.794 vs 0.666).

Because imaging cost is a key constraint, we evaluated modality-level contributions and found that only cardiac long-axis cine MRI and aortic distensibility MRI contributed substantial independent value. After adjusting for baseline traits, liver, pancreas and DXA features added no significant predictive power. We constructed a joint time-to-event model integrating imaging, a polygenic risk score (PRS) trained on more than 1.25 million individuals from non-imaged UK Biobank participants, the Million Veteran Program and FinnGen, blood biochemistry and clinical variables. The joint model achieved a C-index of 0.75, exceeding PREVENT (0.71) and Framingham Risk Score (0.66). Importantly we found that imaging and genetic risks were largely independent, indicating that individuals with similar imaging-based risk at a given age may progress differently based on underlying genetic risk. A hierarchical risk stratification framework combining clinical, genetic and imaging data identified a subgroup with a 15-fold increase in incident CAD risk relative to a low-risk baseline. Performance was consistent across data collection centers spanning rural and urban settings and diverse demographics (C-index range 0.73–0.76). Our findings demonstrate the utility of multi-modal AI for medical forecasting of common complex disease to preempt or mitigate their occurrence.

## Introduction

A key public health priority is the early identification of individuals at high risk for CAD to enable enhanced screening and targeted preventive therapies. CAD remains the leading cause of death worldwide, accounting for around 18 million deaths each year^1^. Despite therapeutic advances, atherosclerotic plaques often progress silently for decades and nearly half of patients first present with CAD only after an acute myocardial infarction or other major event^2^.

For more than four decades, risk prediction in primary care has relied on statistical equations that aggregate a small set of demographic and laboratory variables. The Framingham Risk Score^3^ and its successors the Pooled Cohort Equations^4^ and PREVENT equations^5,6^ in the United States, QRISK3^7^ in the United Kingdom and SCORE2^8^ in Europe use age, sex, blood pressure, lipids, medication history and smoking status to estimate 10-year risk. Although widely implemented, these models systematically under-or over-estimate risk in individuals and cannot capture the substantial genetic and subclinical disease burden that accrues before symptoms appear.

Family, twin and adoption studies attribute roughly 40-60 % of CAD susceptibility to inherited factors^9^. Genome-wide association studies have mapped hundreds of common variants associated with CAD^10,11^, and aggregating these into genome-wide polygenic risk scores (PRS) has identified individuals whose lifelong risk rivals that of monogenic disorders^12^. Multi-ancestry PRS now incorporate data from more than one million individuals and further improve discrimination across different populations^13^. However, genetic risk prediction is ultimately limited by heritability, and it has been shown that lifestyle modification can work in concert with genetic risk^14^, underscoring the value of integrating environmental risk.

Non-invasive imaging can capture a proportion of the genetic risk but also environmental risk by directly visualizing atherosclerotic plaque, cardiac remodeling, body fat and other factors known to influence adverse CAD outcomes. A variety of imaging modalities, including coronary computed tomography angiography (CCTA), positron emission tomography (PET), carotid ultrasound, and cardiac MRI, have been employed to derive these biomarkers^15–21^. Several groups have utilized deep learning models to quantify these factors^22–29^. For example, coronary computed tomography, derived perivascular fat attenuation index has been used to quantify coronary inflammation and predict cardiac mortality independent of calcium score^30^. In addition, population-scale cardiac magnetic-resonance phenotyping has yielded automated measurements of ventricular structure, function and aortic elasticity^31^, extracted from deep learning approaches^32^. Imaging-wide association studies have also been conducted to examine the genetic basis of cardiac morphology and function^33^.

In addition to genetic and imaging data, large biobanks also provide information from other high-dimensional omics data, such as plasma proteomics, offering additional predictive information beyond traditional risk factors. For example, a recent study showed that a panel measuring approximately 5,000 plasma proteins provided clinically meaningful improvement in risk reclassification over both polygenic risk scores (PRS) and conventional clinical variables^34^.

However, datasets presenting combined, imaging, genetic, metabolic and health record data have only become available at the scale necessary to utilize them for prediction until recently and no study to date has systematically combined raw multi-organ imaging, genome-wide genetic information, metabolomic, and demographic data into a unified framework for CAD risk prediction^35^. Existing approaches while outperforming traditional scores^36–38^ typically rely on derived imaging traits, conventional risk factors, or partial omics integration, limiting their ability to fully capture the biological heterogeneity underlying CAD onset.

To address this gap, in this study, we developed *preCog*, a scalable and interpretable multimodal AI framework that integrates imaging data from cardiac, hepatic, and vascular systems with genome-wide PRS, metabolomic profiles, and demographic features. Our objectives were threefold: (1) to quantify the predictive value contributed by each modality, including understanding whether all imaging modalities contribute significantly to prediction (2) to explore interactions between genetic risk and imaging-derived features, and (3) to evaluate whether multimodal integration improves incident CAD prediction beyond current clinical standards. By leveraging recent advances in computer vision, survival modeling, and machine learning, we aim to create a unified, biologically informed approach to proactive cardiovascular risk stratification.

## Results

### A Unified Multimodal Framework for Predicting 10-Year CAD Risk

To implement this framework, we trained *preCog*, a multimodal AI model integrating cardiac imaging, body composition imaging, genetic risk, metabolic biomarkers, ECG features, and demographic data to predict 10-year CAD risk (workflow illustrated in **Fig. 1**). We leveraged the UK Biobank (UKB)^39,40^ which offers a uniquely comprehensive resource combining longitudinal baseline, genetic, blood biochemistry, imaging, and health outcome data.

**Figure 1:**
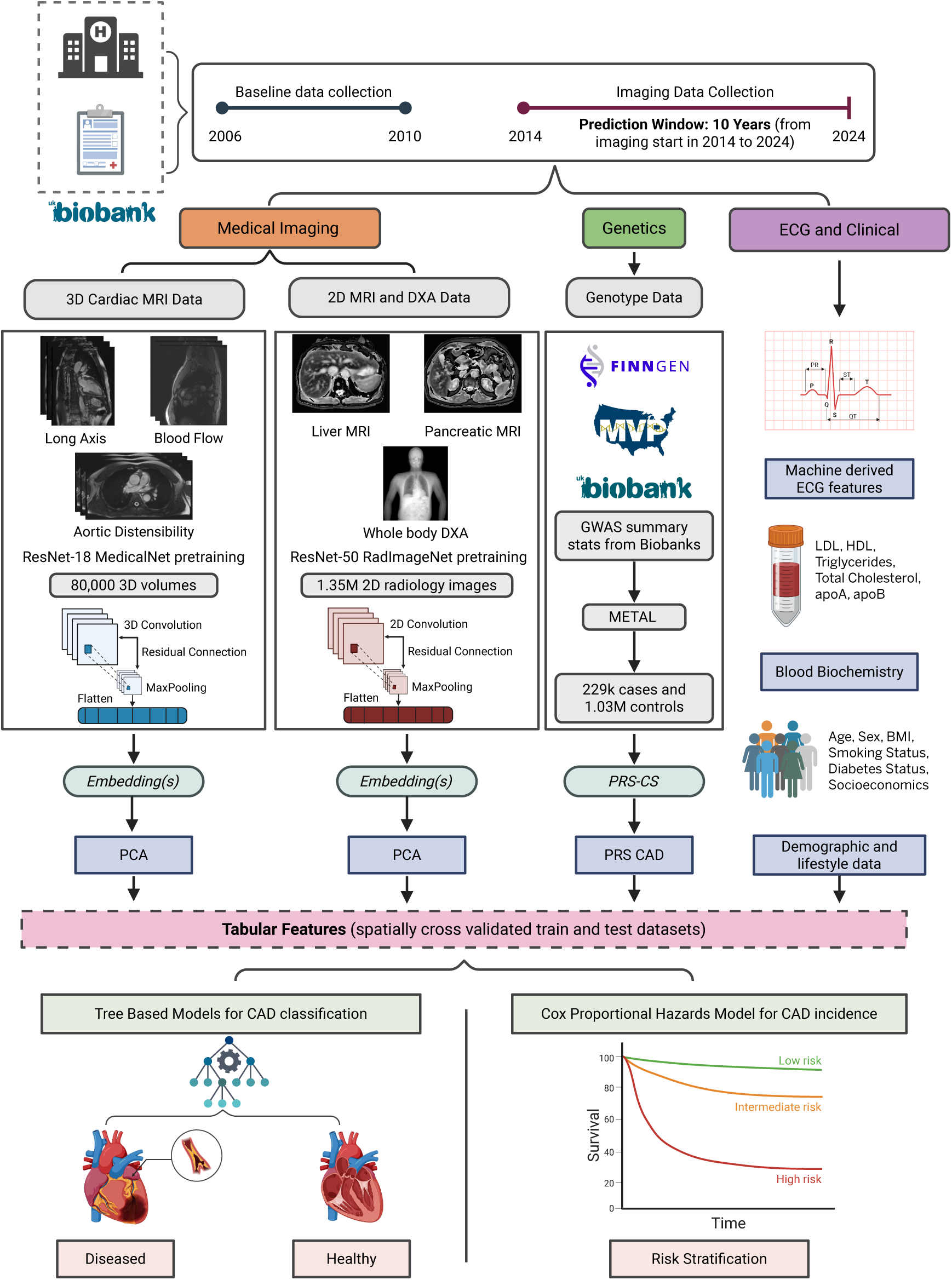
Overview of the *preCog* multimodal risk prediction framework for CAD. The UKB dataset was used to develop a personalized 10-year CAD risk model integrating medical imaging, genetic, metabolic, and demographic data. Baseline data collection occurred between 2006 and 2010, with imaging visits starting in 2014, enabling prospective risk modeling through 2024. **Left:** Imaging data include 3D Cardiac MRI (long-axis views, blood flow, aortic distensibility) processed using a fine-tuned MedicalNet^42^ ResNet-18 model, and 2D imaging (liver and pancreas MRI, and DXA) processed using a ResNet-50 model pretrained on RadImageNet^47^. Embeddings extracted from each modality were reduced using PCA for downstream integration. **Middle:** Genetic datasets were leveraged to compute a PRS (using PRS-CS^49^) for CAD, derived from multi-ancestry GWAS summary statistics across UKB^39,40^, FinnGen^50^, and the Million Veteran Program (MVP)^51^. **Right:** ECG time-series were converted into structured features capturing cardiac electrophysiology, and blood-based metabolic biomarkers (LDL, HDL, triglycerides, total cholesterol, apoA, and apoB) were extracted alongside demographic and lifestyle information (age, sex, BMI, smoking, diabetes status, and socioeconomic indicators). All features were combined into a tabular dataset for spatially cross-validated training and testing. Two downstream models were developed: Tree based classifiers for binary CAD classification, and a Cox proportional hazards model for time-to-event analysis.

Between 2006 and 2010, UKB enrolled over 500,000 participants, collecting comprehensive baseline data including demographics, lifestyle factors, metabolic biomarkers, and genome-wide genotyping. In 2014, UKB launched a large-scale imaging enhancement project that continues to the present, capturing multimodal imaging across four sites Bristol, Cheadle, Newcastle, and Reading encompassing cardiac MRI, abdominal MRI of the liver and pancreas, and whole-body DXA scans. For this study, we analyzed each participant’s first imaging visit and defined incident CAD based on clinical diagnoses (see **Definition of CAD**, in Methods) recorded after this timepoint, allowing a prospective follow-up period of up to 10 years, through 2024. Notably, our outcome phenotype is composed primarily of symptomatic disease events, such as myocardial infarction and angina (including death due to these), while our imaging data captures subclinical cardiovascular features. Thus, our model effectively learns to predict which individuals with early, asymptomatic signs of atherosclerosis are most likely to progress to overt and clinically recognized CAD.

The imaging data were processed using foundational models for vision-based deep learning pipelines tailored to each modality. Cardiac MRI including long-axis views, blood flow, and aortic distensibility sequences was encoded using 3D convolutional neural networks pretrained on large-scale medical imaging datasets. In parallel, liver and pancreas MRIs as well as whole-body DXA scans were embedded using 2D convolutional models. For each modality, high-dimensional embeddings were compressed via principal component analysis (PCA)^41^, retaining the most informative components while reducing dimensionality (see **3D Cardiac MRI Data and Model** and **2D MRI DXA Data and Models**, in Methods).

These imaging-derived principal components were integrated with additional risk factors, including genetic risk quantified by a custom multi-ancestry CAD PRS. This PRS outperformed GPS_mult_^13^ and a UKB-specific case-control PRS (Cox and Snell R² = 0.0217 vs. 0.0194 vs.

0.0187; **Supplementary Fig. 6**; see **GWAS and PRS Construction and Evaluation** in Methods). We further incorporated ECG-derived conduction and waveform features (e.g., PR interval, QRS duration, T-wave amplitude), serum metabolic biomarkers (e.g., LDL, HDL, total cholesterol, triglycerides), and demographic traits (e.g., age, sex, BMI, smoking, medication use, socioeconomic status). (**Supplementary Tables 6–7**; see **Electrocardiogram data** and **Metabolic and Demographic Data** in Methods).

We then combined all the features into a single model and using tree-based algorithms for CAD classification and Cox proportional hazards models for time-to-event prediction. Imaging, genetic and clinical variable preprocessing were carried out using standardized pipelines for each data modality (see **Multimodal Risk Modeling and Feature Selection** in Methods), and generalizability was assessed through spatial cross-validation across imaging centers (**Fig. 1**).

### Vision model training and optimization

To quantify imaging based CAD risk, we constructed two training datasets to evaluate the impact of class imbalance on model performance: one reflecting the natural imbalance in the population (4000 CAD cases and 53,476 controls), and another balanced dataset consisting of 4,000 CAD cases and 4,000 controls. Models trained on the imbalanced dataset achieved an AUC of 0.664 but exhibited poor calibration, with a high false positive rate (FPR = 0.538) and low precision (0.076) (**Supplementary Fig. 1;** see **Data Partitioning and Initial Imbalanced Modelling** in Methods). In contrast, training on the balanced dataset improved overall discrimination, yielding an AUC of 0.700 and accuracy of 0.678 (**Supplementary Fig. 2**). Given these improvements, we adopted the balanced dataset for all downstream model training to mitigate overfitting to the majority class and ensure better generalizability. An independent test set of 200 incident CAD cases and 200 matched controls, and another imbalanced test set of 200 incident cases and 3,000 controls was held out for final evaluation (see **Final Dataset Construction for Imaging Models** in Methods).

For cardiac imaging, we extracted latent feature embeddings from three different data modalities: long-axis cardiac MRI, blood flow MRI, and aortic distensibility MRI using fine-tuned 3D convolutional networks^42–45^. We examined the performance of several types of pretraining datasets (including 3d action recognition and 3d medical imaging datasets) for our task of disease risk prediction. Amongst these competed pretraining models, models initialized with MedicalNet^42^ pretraining demonstrated superior performance (AUC = 0.700, accuracy = 0.678) relative to other video-pretrained counterparts^43,44,46^ (**Supplementary Fig. 2 and Supplementary Table 2**; see **3D Cardiac MRI Data and Model** in Methods). To capture broader cardiometabolic risk encompassing other organs, we integrated features from 2D liver and pancreas MRI and whole-body DXA scans. We again compared the performance of different pretrained models and found that models trained on RadImageNet (a large-scale medical imaging dataset comprising over 1.3 million annotated CT, MRI, and ultrasound scans across a variety of anatomical regions)^47^ on top of a ResNet-50 backbone^45^, improved CAD classification compared to a standard ImageNet-pretrained models^48^ (**Supplementary Fig. 3** and **Supplementary Table 3**; see **2D MRI, DXA Data and Models** in Methods).

To evaluate the informativeness of the learned representations from each imaging model, we first applied principal component analysis (PCA)^41^ to the raw embeddings extracted from the final convolutional layers (see **Embedding Extraction, Dimensionality Reduction, and Benchmarking** in Methods). Across views, the first two principal components captured over 70 percent of the variance, and by ten components, models achieved nearly the same performance as with the full embeddings (**Fig. 2a** and **Supplementary Table 4**). These low-dimensional representations offered consistent separation between CAD cases and controls, particularly along the first principal axis (**Supplementary Fig. 4**). This suggests that deep networks can compress large-scale spatiotemporal image data into compact, task-relevant manifolds.

**Figure 2:**
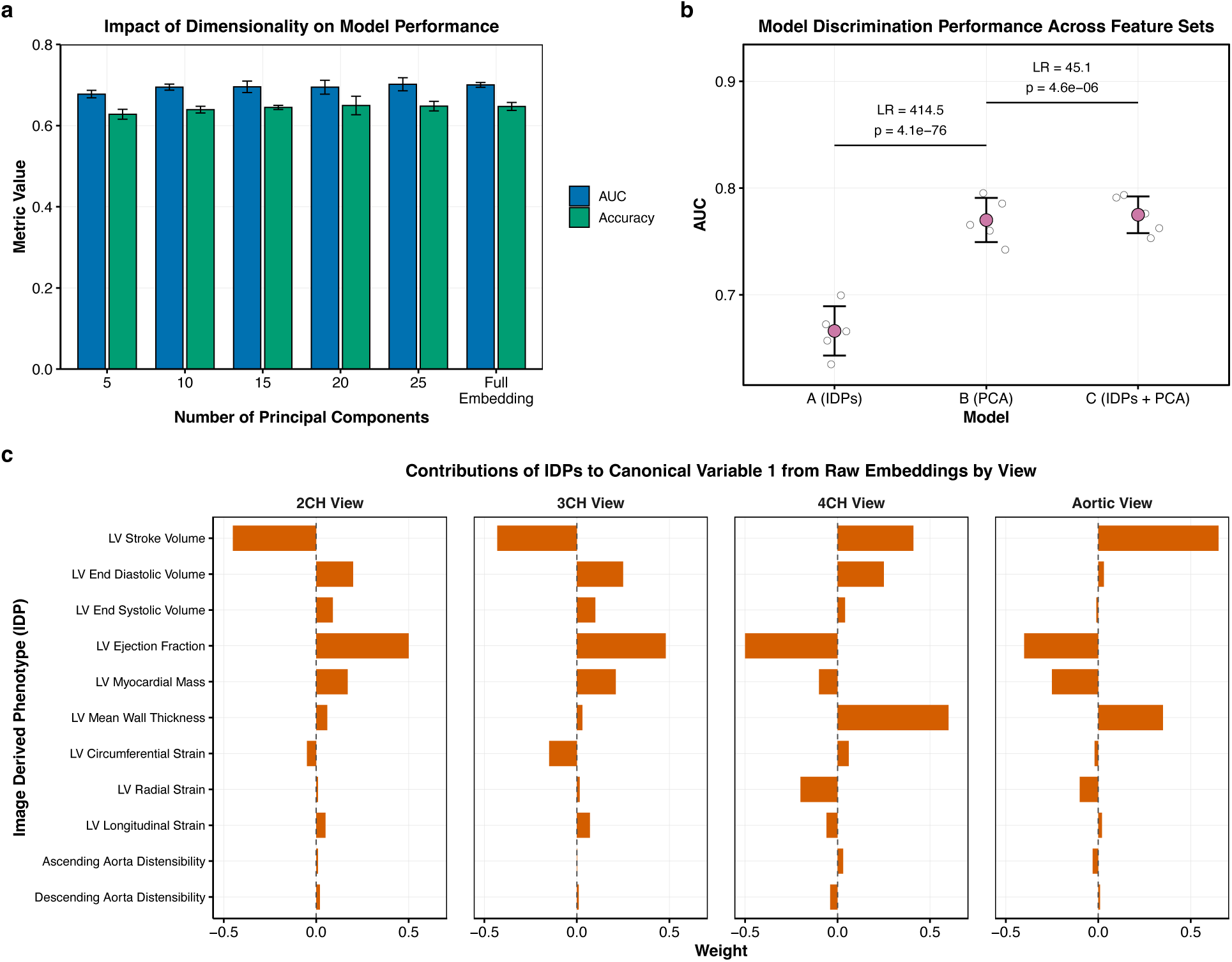
Dimensionality reduction, predictive performance, and biological interpretability of deep learning-derived embeddings. **a**, Performance of logistic regression models trained on increasing numbers of principal components (PCs) extracted from imaging embeddings, showing both AUC and accuracy across five-fold cross-validation. As few as 10 PCs per modality retain most of the predictive power, with diminishing gains beyond 25 PCs. **b**, Comparison of AUC across three feature sets: traditional imaging-derived phenotypes (IDPs), PCA-reduced deep embeddings, and their combination. PCA-based features substantially outperform IDPs (*p* < 1e-75, likelihood ratio test), while adding IDPs offers only a marginal improvement (*p* < 1e-5), suggesting that learned representations capture most of the signal available in handcrafted traits. **c**, Canonical correlation analysis (CCA) of raw embeddings from four cardiac MRI views (2, 3 and 4 chamber views from cardiac long axis MRI and cine aortic distensibility MRI) with matched IDPs, revealing which phenotypes dominate the learned representation. Traits such as stroke volume, ejection fraction, and myocardial mass consistently show high loadings on the first canonical variable, indicating that these biologically relevant and visually prominent features are strongly embedded in the model’s latent space.

To benchmark these PCA-reduced imaging embeddings, we compared them to conventional imaging-derived phenotypes (IDPs) such as volumes, mass, and strain from expert-annotated segmentation pipelines. PCA-based models achieved a higher area under the receiver operating characteristic curve (AUC = 0.794) than IDP-based models (AUC = 0.666), while combining both yielded only a modest gain (AUC = 0.799; **Fig. 2b** and **Supplementary Table 5**).

Likelihood ratio tests confirmed that PCA features captured the bulk of predictive information (LR = 414.5, *p* < 1e-75), with minimal added benefit from IDPs (LR = 45.1, *p* < 1e-5). These findings indicate that automatically learned features can substitute for traditional phenotypes, with advantages in scalability and dimensionality.

To interpret what the embeddings encode, we performed canonical correlation analysis (CCA)^52^ between raw embeddings from long axis and aortic distensibility MRI and IDPs derived from the same view. CCA identifies linear combinations of features from both sets that are maximally correlated, revealing which biological traits are reflected in the learned representations. The first canonical variable from each MRI view was strongly aligned with a small number of cardiac IDPs, particularly stroke volume, ejection fraction, myocardial mass, and cavity volumes (**Fig. 2c, Supplementary Figure 5 and Embedding Extraction, Dimensionality Reduction, and Benchmarking** in Methods). Thus, while embeddings achieved superior predictive performance compared to IDPs, their most heavily weighted dimensions nonetheless recapitulated phenotypes known to affect cardiovascular disease risk. This dual property underscores that embeddings function as task-optimized generalizations of IDPs: they capture the discriminative signal of expert-derived traits while extending beyond them to encode additional, subtler patterns.

The traits with the highest canonical loadings are also those most tightly linked to CAD pathogenesis and its downstream effects. Increased myocardial mass and wall thickness are hallmarks of hypertensive and ischemic remodeling and predict future CAD and heart failure events^53,54^. End-diastolic and end-systolic volumes increase in response to dilatation and impaired contractility, which are common in subclinical ischemia^55^. Stroke volume and ejection fraction, as integrated measures of systolic function, reflect infarct burden and ventricular stunning and are established correlates of CAD severity^56,57^. By contrast, features such as global strain and aortic distensibility were weighted lower, likely due to their reduced measurement fidelity or more subtle signal in these networks. Together, these results suggest that deep embeddings recapitulate precisely those physiologic traits that are not only visually prominent in MRI but also mechanistically and prognostically linked to coronary disease.

### Genetic, ECG, Metabolic, and Demographic Feature Processing

To complement the imaging modalities, we incorporated a comprehensive set of non-imaging features spanning genetic, electrophysiological, metabolic, and demographic domains. For quantifying inherited risk, we constructed a custom PRS for CAD. This PRS was derived from meta-analyses of multiple large-scale genome-wide association studies (GWAS), encompassing 229,724 CAD cases and 1,024,752 controls. Specifically, we integrated GWAS summary statistics from three major cohorts: non-imaged White British participants from the UKB, FinnGen^50^, and the Million Veteran Program (MVP)^51^ (**Supplementary Figure 6**; see **GWAS and Polygenic Risk Score (PRS) Construction and Evaluation** in Methods).

Electrocardiographic (ECG) features were extracted from resting 12-lead electrocardiograms using automated interpretation software. These included a panel of machine-captured conduction and waveform metrics, such as PR, QRS, QT, and RR intervals, as well as average P, Q, R, and T wave amplitudes. These features provided complementary information on cardiac electrical function, conduction velocity, and rhythm abnormalities, and offered insight into subclinical changes in cardiovascular physiology (**Supplementary Table 6**; see **Electrocardiogram Data** in Methods).

Metabolic and demographic data were derived from the UKB baseline assessments. Metabolic biomarkers included serum concentrations of LDL, HDL, triglycerides, total cholesterol, apolipoprotein A, and apolipoprotein B, each of which reflects different aspects of lipid metabolism. Demographic covariates included age, sex, body mass index (BMI), smoking status, diabetes mellitus diagnosis, medication history (for insulin, lipid lowering, and blood pressure lowering drugs), household income, and the Townsend deprivation index as a proxy for socioeconomic status (**Supplementary Table 7**; see **Metabolic and Demographic Data** in Methods). All continuous variables were z-score normalized, and categorical variables were one-hot encoded.

### Multimodal Risk Modeling and Feature Selection

To refine CAD risk prediction, we constructed a multimodal dataset combining imaging-derived principal components, PRS, metabolic biomarkers, ECG features, and demographic variables (see **Multimodal Risk Modeling and Feature Selection** in Methods). Each modality was distilled into low-dimensional representations and combined using a late fusion strategy, enabling linear modeling while reducing complexity and overfitting risk. After excluding individuals with missing data, the training set included 3,000 CAD cases and 3,000 controls, with independent balanced (200 cases, 200 controls) and imbalanced (200 cases, 3,000 controls) test sets.

Regularized logistic regression models trained on individual feature groups showed that long-axis cardiac MRI had the highest standalone predictive power (Nagelkerke pseudo R_N_² ≈ 0.22), followed by medication history, aortic distensibility imaging and demographic features (R_N_² > 0.15; **Fig. 3b** and **Supplementary Table 9**). To study the impact of a single predictor in a joint model, we performed leave-one-out analysis to estimate the reduction in R_N_². This identified long-axis imaging (∼13% drop), medication history (∼11% drop), PRS (∼8% drop), and aortic distensibility (∼5% drop) as the most critical contributors (**Fig. 3a** and **Supplementary Table 8**). Commonality analysis revealed two important insights (**Fig. 3c** and **Supplementary Table 10**). First, blood flow, liver, pancreas, and DXA imaging had limited additive value once baseline features were included, underscoring their redundancy. Second, long-axis and aortic distensibility imaging retained substantial unique variance even when combined with other modalities, highlighting their non-redundant contribution. Importantly, PRS preserved unique predictive value even after accounting for imaging and clinical features, indicating that genetic and imaging risk capture largely independent dimensions of CAD susceptibility.

**Figure 3:**
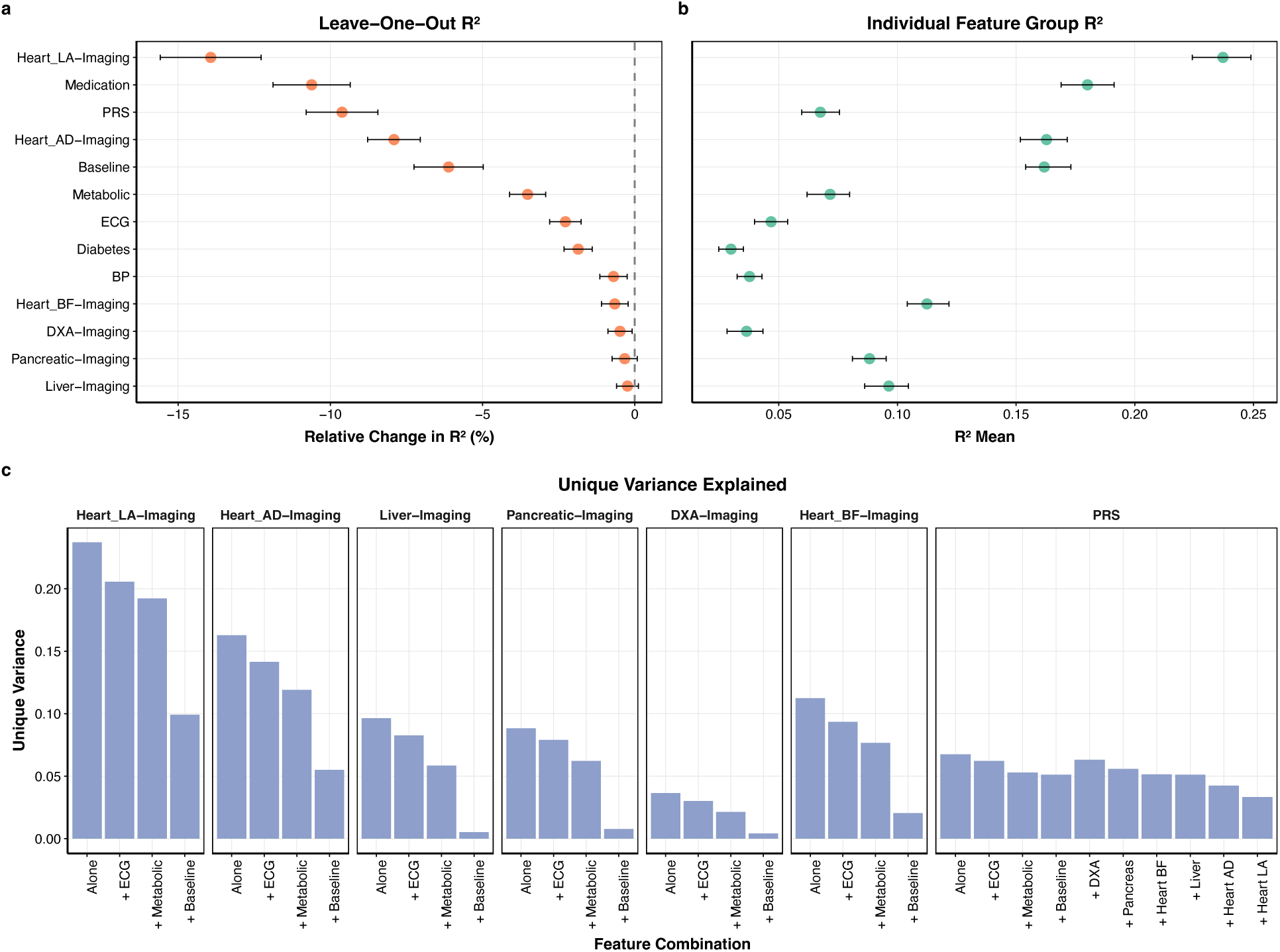
**Contributions of feature groups to CAD prediction**. **a**, Leave-one-out (LOO) analysis showing the relative change in Nagelkerke pseudo R_N_² when individual feature groups are removed from the full multimodal model. Long-axis cardiac MRI, medication history, and PRS drive the largest drops in performance, followed by aortic distensibility imaging and baseline traits (age, sex, BMI, smoking). Liver, pancreatic, DXA, and blood flow imaging contribute minimally. Error bars denote 95% CI. **b,** Standalone performance of models trained on each feature group. Long-axis and aortic distensibility imaging, together with medication history, achieve the highest R_N_², with baseline and metabolic features also showing strong predictive value. Error bars denote 95% CI. **c,** Commonality analysis decomposing explained variance into unique components across feature combinations. Liver, pancreas, DXA, and blood flow imaging lose nearly all unique predictive value once baseline features are included, reflecting redundancy with body composition signals such as BMI. In contrast, long-axis and aortic distensibility imaging preserve substantial unique variance, indicating orthogonal predictive utility. PRS shows modest standalone R_N_² but high unique variance, demonstrating that genetic risk captures information independent of imaging and clinical features.

Based on these analyses, we defined a refined multimodal model incorporating long-axis and aortic distensibility imaging, medication history, PRS, demographics, ECG features, metabolic biomarkers, and diabetes status, while excluding modalities with minimal additive value. This yielded a compact and interpretable architecture (**Supplementary Fig. 7**; **Supplementary Tables 11–13**).

### Enhancing Clinical Risk Prediction with Genetic and Imaging Features

To assess whether genetic and imaging biomarkers improve CAD risk prediction beyond traditional clinical models, we trained nested Cox proportional hazards models and evaluated them on an independent test set (see **Multimodal Survival Modeling and Imaging-Enhanced Risk Stratification** in Methods). Using clinical biomarkers based on Framingham Risk Score (FRS)^58^ components, age, sex, BMI, smoking, lipids, medication use, and diabetes we achieved a test-set concordance index (C-index) of 0.65. Adding PRS increased performance to 0.69, and further inclusion of imaging-derived principal components raised the C-index to 0.75 (**Fig. 4a**). We then benchmarked our model against established tools. On the same test set, the Framingham 10-year risk score and PREVENT^59^ model achieved lower concordance compared to our multimodal model, which provided the highest discriminative power (**Fig. 4b**).

**Figure 4:**
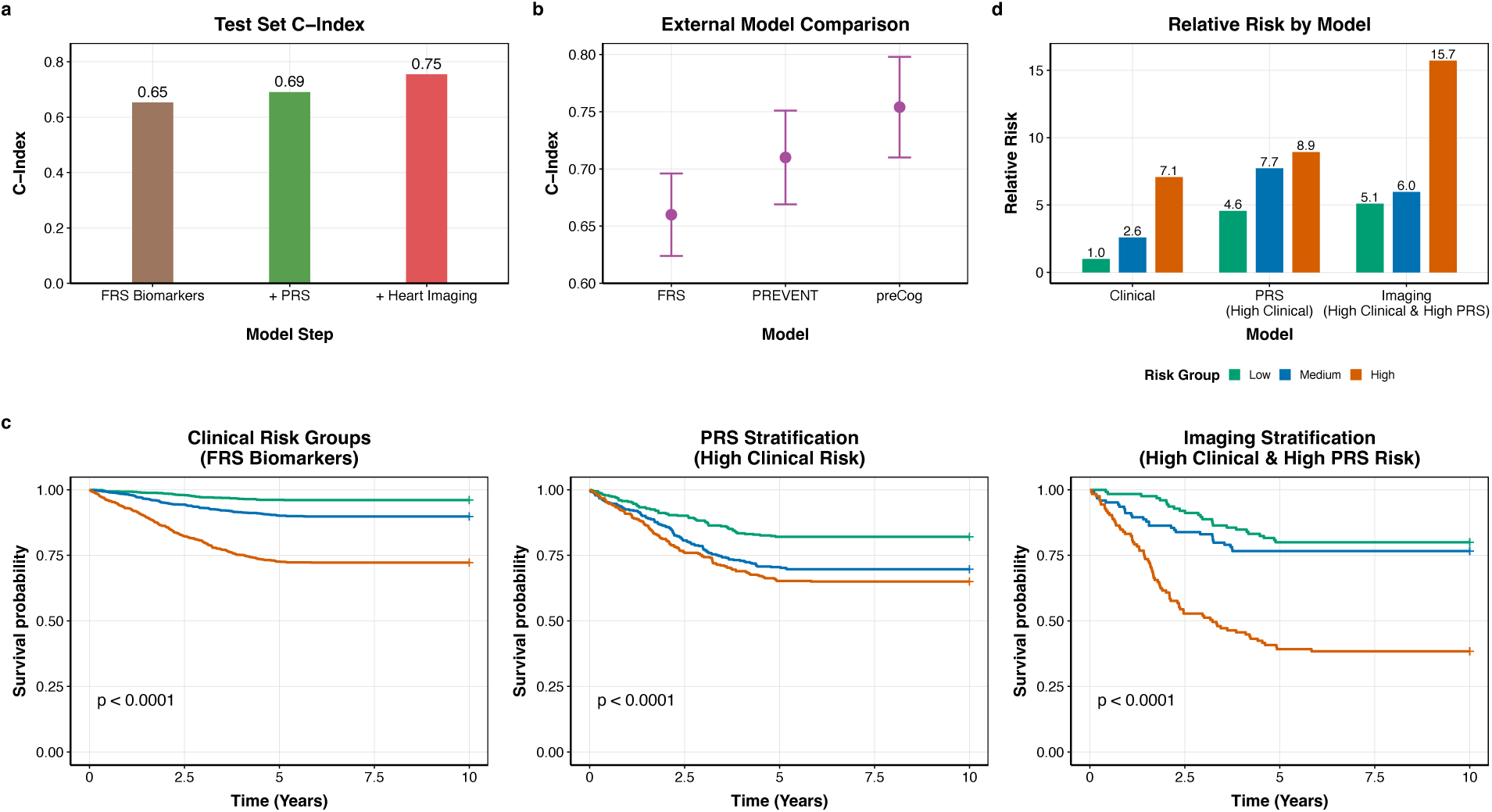
**Multimodal risk modeling improves CAD prediction and enables tiered stratification**. **a**, Test-set C-index for nested Cox models trained sequentially with clinical covariates, and plus PRS, and plus cardiac imaging features. Adding PRS and imaging incrementally improved performance, with the full model achieving the highest C-index (0.75). **b**, External model comparison on the same test set. The Framingham 10-year Risk Score and PREVENT model underperformed relative to our multimodal model (preCog), which showed the highest concordance. **c**, Kaplan–Meier survival curves illustrating stepwise stratification. Left: clinical risk groups defined by FRS biomarkers. Middle: additional stratification of high-clinical-risk individuals using PRS. Right: further stratification of individuals with both high clinical and high PRS using imaging features. Each layer of stratification produced progressively sharper survival separation, with imaging identifying a subgroup at markedly elevated risk. **d**, Relative risk of CAD incidence across models. Clinical features alone identified a high-risk group with RR = 7.1. PRS stratification refined this to RR = 8.9, and imaging revealed a subgroup with RR = 15.7 compared to the baseline low-risk group.

To evaluate risk refinement, we implemented a hierarchical stratification framework. Participants were first stratified into clinical risk groups using clinical biomarkers, followed by PRS stratification within the high clinical-risk group, and finally by imaging features. Kaplan–Meier curves demonstrated progressive separation of survival probabilities across each step, with imaging features providing the strongest discrimination among high-risk individuals (**Fig. 4c**).

Quantitative risk estimation underscored this improvement. The high-risk group defined by clinical features alone had a relative risk (RR) of 7.1. Stratification with PRS increased RR to 8.9, and the addition of imaging features identified individuals with an RR of 15.7 compared to the baseline group (**Fig. 4d**). These results demonstrate that multimodal enrichment uncovers extreme-risk individuals who remain hidden within broader clinical categories.

Taken together, our findings support a cost-effective, tiered risk assessment workflow: apply clinical scoring broadly, integrate genotyping for those at elevated clinical risk, and reserve cardiac imaging for individuals with both high clinical and genetic risk. This stepwise approach optimizes resource allocation while enabling more precise and personalized CAD risk management.

### Robust Generalization Across Imaging Centers via Spatial Cross-Validation

To evaluate the generalizability of our multimodal framework, we leveraged the UKB’s geographic diversity through a spatial cross-validation strategy (**Supplementary Table 14**; see **Spatial Cross-Validation to Evaluate Site-Level Generalizability** in Methods). Imaging data from four distinct centers Bristol, Cheadle, Newcastle, and Reading enabled a leave-one-center-out design, training on three centers and evaluating on the fourth.

Our multimodal classifier achieved stable performance across sites, with AUCs ranging from 0.785 to 0.822 (**Fig. 5b**). Similarly, survival models maintained consistent concordance indices across centers ranging from 0.73 to 0.76 (**Fig. 5c**), confirming robust generalization even amid site-specific differences in scanner hardware, operator protocols, and participant demographics^53^ (**Fig. 5a**).

**Figure 5:**
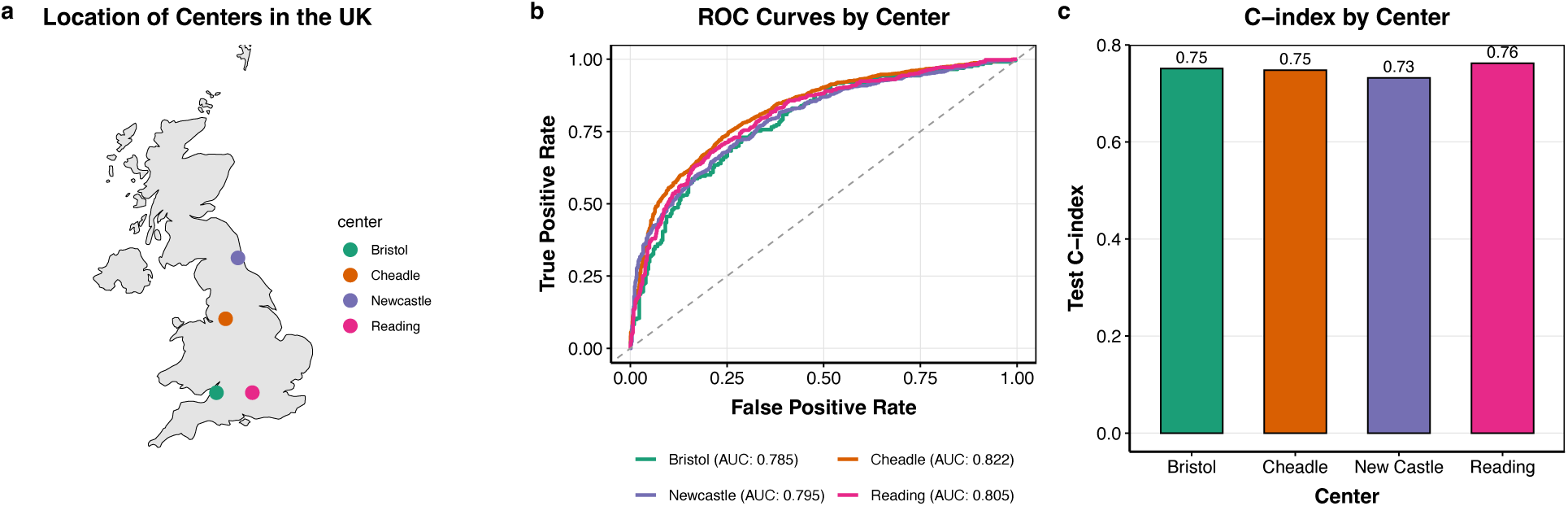
Spatial cross-validation confirms model generalizability across imaging centers. **a**, Geographic distribution of the four UK Biobank imaging centers Bristol, Cheadle, Newcastle, and Reading used for spatial cross-validation. **b**, Receiver operating characteristic (ROC) curves for the multimodal XGBoost model, trained using features from imaging (MedicalNet-extracted principal components), baseline demographics, metabolic biomarkers, and diabetes status. Each curve represents model performance when trained on data from three centers and evaluated on the held out fourth. Area under the curve (AUC) values remain consistent across centers, indicating strong generalization. **c**, C-index for Cox proportional hazards models evaluated in the same leave-one-center-out fashion, demonstrating stable time-to-event prediction across centers. Together, these results show that the full multimodal risk prediction pipeline spanning deep imaging models, feature fusion, and survival analysis maintains robust performance across diverse acquisition environments and populations.

These findings validate the end-to-end robustness of *preCog*’s modeling pipeline from deep imaging feature extraction and dimensionality reduction to multimodal integration and outcome prediction. Despite geographic and technical heterogeneity, *preCog* maintained reliable classification and survival performance.

## Discussion

CAD evolves from subclinical changes to acute clinical events which often result in premature death, making early identification during the asymptomatic phase a key challenge in preventive cardiology. Here, we introduced *preCog*, a multimodal AI framework that integrates imaging-derived features, PRS, metabolic biomarkers, and demographic traits to predict 10-year CAD risk. Our findings demonstrate that *preCog* substantially improves risk prediction accuracy beyond traditional clinical baselines, achieves robust generalization across geographically distinct imaging centers in the UKB, and offers a scalable strategy for personalized cardiovascular care.

Our study has 3 major conclusions. First, our analysis examined the extent to which different imaging modalities provide additional power to discriminate between incident cases and controls. Each type of imaging captures a slightly different part of the overall risk profile. For example, liver fat or abdominal fat could capture some aspects of the metabolic profile of the individual. However, obtaining imaging data from across the body at the population level is prohibitively expensive, and there is a need to identify the type of imaging modalities that might be most predictive of this disorder. Through systematic feature selection and dimensionality reduction, we found that long-axis cardiac MRI and aortic distensibility MRI features were the only two imaging modes that were independently predictive when combined with other features suggesting that cardiac imaging alone is sufficient for disease prediction at least to first order.

This finding is consistent with clinical evidence, as both left ventricular function and aortic stiffness are recognized as early markers of CAD^60,61^. Our work also showed the value of direct encoding of image risk using deep learning-derived principal components instead of conventional IDPs in risk prediction. This approach also had the added advantage of maintaining compact, scalable representations for use in settings with relatively small sample sizes.

In addition, we found that imaging risk and genetic risk were relatively complementary, suggesting that perhaps disease progression in individuals presenting similarly with imaging at a particular age might differ based on an individual’s genetic profile. These insights, obtained through leave-one-out and commonality analyses, underscore the importance of prioritizing structural and vascular cardiac imaging alongside genetic risk profiling in CAD modeling. This suggests that individuals who are at high genetic risk might need to be imaged more frequently as compared with people who do not over the course of their lifetime to achieve similar rates of prevention.

Finally, our study also demonstrated the power of a multi-modal approach for a major disease prediction task. Augmenting traditional clinical models with genetic and imaging features led to substantial gains in predictive performance. Integrating PRS and imaging-derived principal components into clinical biomarkers based Cox models increased the test C-index from 0.65 to 0.75, and hierarchical stratification strategies highlighted individuals with more than fifteen-fold elevated risk who would otherwise be masked within broad clinical categories. These findings support a pragmatic, cost-effective tiered workflow: initial screening with clinical risk scores, followed by selective genotyping, and targeted cardiac imaging for those with combined high clinical and genetic risk.

Despite the promise of *preCog*, limitations remain. Cardiac MRI, while offering rich structural and functional insights, is not a frontline modality in routine clinical CAD evaluation as it only captures some of the proximal coronary arteries, however it is able to capture cardiac structure and function^60,62^. Extension to more widely available modalities, such as coronary computed tomography angiography (CCTA) or echocardiography, will be critical for broader clinical applicability. Furthermore, although spatial cross-validation across UKB imaging centers provided rigorous internal validation, external validation across independent healthcare systems and more ancestrally diverse populations is essential to establish generalizability.

In conclusion, our work demonstrates that multimodal AI integrating imaging, genetic, metabolic, and clinical data can substantially advance early CAD risk prediction. By coupling deep phenotyping with a scalable deployment strategy, *preCog* represents a significant step toward precision cardiology. Future efforts should focus on adapting this framework to routine clinical imaging, expanding validation across global populations, and collaborating with health systems to support real-world clinical implementation.

## Methods

### Data Source

The UK Biobank (UKB) is a large-scale, population-based study conducted in the UK, enrolling around 500,000 adults aged 40–69 between 2006 and 2010. Designed to explore genetic and lifestyle influences on a wide range of diseases, the UKB provides an extensive dataset, including baseline information, genomic data, and Electronic Health Records (EHR) for half a million individuals^39,40^. Additionally, UKB includes medical imaging data, covering MRI scans (for the heart, liver, and pancreas), ECGs, and DXA scans (with whole-body fat composition) for over 65,000 individuals.

For this study, we compiled multimodal data from 80,000 individuals with a CAD prevalence of approximately 7%. In our survival models, we used data from the first imaging visit (2014–2024) as the incidence prediction window, excluding cases that occurred prior to this period. Data access for this study was granted under UKB application ID 65439.

### Definition of CAD

CAD, the primary outcome of this study, was defined using diagnostic and procedural codes recorded in participants’ electronic health records (EHR). Diagnosis of CAD was based on a combination of ICD-10, ICD-9, and OPCS-4 codes indicative of ischemic heart disease or related revascularization procedures.

From the ICD-10 classification, we included codes representing chronic ischemic heart disease and acute myocardial infarction, as well as angina and related sequelae. Specifically, the following codes were used: I20–I25 (covering angina pectoris, acute myocardial infarction, and other forms of chronic ischemic heart disease), including subcodes such as I21.0–I21.9, I22.0– I22.9, I23.0–I23.8, I24.0–I24.9, I25.0–I25.9, I25.2, Z95.1 (presence of aortocoronary bypass graft), and Z95.5 (presence of coronary angioplasty implant and graft). Collectively, these codes capture both acute and chronic manifestations of CAD as well as prior surgical history related to coronary revascularization.

Historical diagnoses from the ICD-9 coding system were also included. We identified CAD using the following codes: 410.9 (acute myocardial infarction), 412.9 (old myocardial infarction), and 414.0, 414.1, 414.8, 414.9 (chronic ischemic heart disease and related subtypes).

Procedural evidence for CAD was captured using OPCS-4 codes related to percutaneous coronary intervention (PCI) and coronary artery bypass grafting (CABG). Included codes span a broad range of surgical interventions involving coronary arteries, specifically: K40–K46 (procedures involving bypass of coronary arteries), K49–K50 (other therapeutic transluminal operations on coronary artery), and K75.1–K75.9 (percutaneous transluminal balloon angioplasty and insertion of stents). Additional related subcodes such as K41*, K42*, K43*, K44*, K45*, K46*, K49*, and K50* were included to comprehensively capture variations in procedural coding across sites and years.

The date of CAD onset was defined as the earliest date of any qualifying diagnostic or procedural code. In cases where CAD was recorded through self-report (e.g., participant responses indicating prior myocardial infarction or coronary intervention), the self-reported age of onset was used to assign an approximate date of diagnosis.

For time-to-event analyses, participants with prevalent CAD those with an event occurring prior to their imaging date were not included.

### Data Partitioning and Initial Imbalanced Modeling

The initial joint imaging cohort consisted of 55,000 controls and 4,000 CAD cases derived from UKB with paired imaging data and long-term follow-up. In our early experiments, we investigated whether training on the full, naturally imbalanced dataset would be sufficient for CAD classification using deep learning. For this, we selected 4,000 CAD cases and a randomly sampled subset of 53,476 controls to fine-tune a 3D ResNet-18^43^ model pretrained on the Kinetics-400 video dataset^46^. The model was trained to classify binary CAD status using aortic distensibility MRI volumes (FID: 20210, 78,684 participants) as input.

As shown in **Supplementary Fig. 1**, the model trained on the imbalanced dataset demonstrated high recall (0.685), indicating that it successfully identified many CAD cases. However, this came at the expense of extremely low precision (0.076) and F1 score (0.137), reflecting a high false positive rate. The confusion matrix revealed that the model tended to classify most individuals regardless of true status as CAD-positive. This pattern suggests that the model had overfit to the majority signal associated with CAD cases in the training set and failed to learn class-discriminative features for non-CAD controls. To mitigate this, we also experimented with alternative loss functions, including focal loss and class-weighted cross-entropy. However, performance remained unchanged, reinforcing that the imbalance itself not just the loss formulation was the key limiting factor.

The implications of these performance deficits are significant. In a clinical risk prediction setting, high recall but poor precision leads to an excess of false alarms, which could result in unnecessary diagnostic workups and resource strain. Moreover, the model’s poor calibration and bias toward the minority class undermine its utility for stratified risk modeling, where reliable differentiation between high-and low-risk individuals is critical. These findings reinforced the need to rebalance the training data to enable the model to learn a more generalizable decision boundary.

### Final Dataset Construction for Imaging Models

Based on these findings, we opted to construct a balanced training dataset by age-matching and under-sampling controls. The final training set included 4,000 CAD cases and 4,000 controls, selected to maintain demographic balance while mitigating the bias observed in the imbalanced setting. In addition, we set aside an independent test set comprising 200 incident CAD cases and 200 controls, where all cases had CAD diagnoses occurring after the imaging date. This test set was used for downstream model evaluation across both classification and survival analysis pipelines. To assess generalizability under real-world prevalence, we also evaluated all models on a separate imbalanced test set consisting of 200 incident CAD cases and 3,000 controls.

### 3D Cardiac MRI Data and Model

Cardiac MRI provides comprehensive information on both the structural and functional aspects of the heart, capturing measures relevant to cardiovascular disease, such as left ventricular mass, left ventricular ejection fraction, left atrial volume, and aortic stiffness. The cardiac MRI scans were performed using a Siemens 1.5 Tesla MAGNETOM Aera scanner (Siemens Healthineers, Erlangen, Germany) equipped with VD13A software and a spine and body flex matrix coil. No pharmacological stressors or contrast agents were used in these scans.

For this study, we utilized Cardiac MRI data in three specific modes. The first mode is the long-axis view (FID: 20208, 83,542 participants), which provides three distinct chamber views of the heart’s long axis: 2-chamber, 3-chamber, and 4-chamber, capturing critical structural features. The second mode is Blood Flow MRI (FID: 20213, 83,752 participants), which reveals blood flow patterns within the heart, essential for assessing both structural and functional dynamics.

The third mode is Aortic Distensibility MRI (FID: 20210, 78,684 participants), which evaluates the elasticity of the aortic wall by measuring changes in aortic diameter throughout the cardiac cycle. Aortic distensibility is a key marker of vascular health, providing insights into arterial stiffness, which has been linked to hypertension, cardiovascular disease, and overall cardiovascular risk^63^.

In processing the long-axis view images, we created a numpy (.npy) file for each individual with a shape of (50, 200, 200, 3), where 50 frames (each 200×200 in resolution) captured the three distinct chamber views (2-chamber, 3-chamber, and 4-chamber) of the heart’s long axis. For Blood Flow MRI, we processed three distinct views: one visualizing blood flow, another indicating flow phase (velocity and direction), and a third capturing flow magnitude. After processing, we saved each individual’s Blood Flow MRI data as a numpy (.npy) file with a shape of (30, 200, 200), ensuring the retention of essential flow and structural information. For Aortic Distensibility MRI, which evaluates the elasticity of the aortic wall throughout the cardiac cycle, a continuous sequence of 100 frames was captured during imaging. The raw DICOM images were preprocessed and converted into numpy (.npy) files with a shape of (100, 200, 200) for each individual.

For CAD prediction from 3D cardiac MRI data, we fine-tuned a suite of video-pretrained and medically-pretrained deep learning models. These included 3D ResNet-18^43^, MC3-18^44^, and R(2+1)D^43^ models pretrained on the Kinetics-400 dataset, as well as a Med3D model pretrained on a large corpus of 3D medical scans^42^. Kinetics-400 pretraining enabled models to learn spatiotemporal patterns from natural video sequences^46^, while Med3D offered domain-specific initialization tailored to volumetric medical imaging tasks.

To match model input requirements, we resized our original MRI datasets. Long-axis cardiac MRI, originally of shape (50, 200, 200, 3), was resized to (16, 112, 112) for each of the three views—2-chamber, 3-chamber, and 4-chamber, effectively capturing cardiac cycle dynamics while balancing memory efficiency. Blood Flow MRI was down sampled from (30, 200, 200) to (16, 112, 112), preserving key hemodynamic features. Aortic Distensibility MRI was similarly reshaped from (100, 200, 200) to (16, 112, 112), ensuring consistent temporal representation of aortic elasticity across modalities.

We trained five independent models, one per imaging subtype, allowing each to specialize in its modality. The three chamber views capture orthogonal anatomical perspectives relevant to CAD, such as regional wall motion, valve dynamics, and chamber dilation. Blood Flow and Aortic Distensibility models further contribute vascular and hemodynamic features crucial for CAD detection. A schematic of the fine-tuning process is illustrated in **Supplementary Fig. 2a.**

All models were fine-tuned using the FastAI framework^64^. We employed a two-stage training strategy using the fine_tune() method: initially training only the classification head, followed by gradual unfreezing and fine-tuning of deeper layers. This approach allowed models to adapt learned representations to CAD-specific features. To improve generalization, we applied data augmentation (random rotations, flipping, brightness variation) and regularization techniques such as weight decay.

As shown in **Supplementary Fig. 2b**, the Med3D model outperformed others across both metrics achieving an AUC of 0.700 and accuracy of 0.678. Among Kinetics-pretrained architectures, R(2+1)D yielded the highest performance (AUC = 0.650, accuracy = 0.639), followed by MC3-18 and 3D ResNet-18 (see **Supplementary Table 2**). These results highlight the added value of domain-specific pre-training for 3D medical image classification and demonstrate the complementarity of spatiotemporal modeling strategies across modalities.

Full training configurations, hyperparameter settings, and reproducible code are available in the GitHub repository linked to this manuscript.

To elucidate the spatial focus of our trained models and gain interpretability into their classification process, we implemented Gradient-weighted Class Activation Mapping (Grad-CAM)^65^. This technique generates heat maps that highlight image regions most influential in the model’s predictions, offering insights into the learned decision-making process. We applied Grad-CAM to the fine-tuned 3D ResNet-18 and Med3D models across three imaging modalities: long-axis cardiac MRI, Blood Flow, and Aortic Distensibility MRI.

Focusing on the ResNet-18 and Med3D models, we extracted feature maps from its second, third, and fourth residual layers (layer 2, layer 3, layer 4) and computed class activation maps using gradients of the predicted class with respect to each feature map. These weighted activations were then visualized on individual MRI slices, specifically from the mid-cardiac cycle (slices 17 to 32), where the myocardium is most structurally informative.

As illustrated in **Supplementary Fig. 2c**, the Grad-CAM overlay for slice 18 (layer 4) of a CAD-positive individual reveals strong activations around the left ventricular myocardium, interventricular septum, and basal segments regions clinically associated with ischemic remodeling. Notably, there is also activation near the right ventricular insertion point, which may indicate secondary remodeling or wall motion irregularities. In contrast, the corresponding Grad-CAM for a control subject displays diffuse and less localized activations, suggesting that the model’s attention is more spatially constrained and diagnostically meaningful in CAD cases.

Layer-wise comparisons further show a progressive refinement of spatial focus across the network’s depth. Earlier layers (e.g., layer 2) capture broader anatomical outlines, while deeper layers (e.g., layer 4) localize sharply to pathophysiologically relevant areas. This trend was consistent across both ResNet-18 and Med3D models, though Med3D often showed slightly broader focus zones, potentially reflecting the influence of its diverse medical pretraining.

By applying Grad-CAM to both CAD cases and controls, we confirmed that the models consistently focused on clinically meaningful regions in affected individuals while showing less structured attention patterns in controls. These interpretable visualizations validate that the models are not relying on spurious correlations or imaging artifacts but are instead capturing biologically plausible features.

Incorporating Grad-CAM into our analysis pipeline not only enhanced transparency but also reinforced the utility of long-axis cardiac and vascular MRI in identifying morpho-functional correlates of CAD. This step toward explainability ensures our models align with clinical intuition, a crucial prerequisite for real-world deployment in risk stratification and diagnostic support.

### 2D MRI, DXA Data and Models

While conventional anthropometric indicators like BMI, weight, and waist-to-hip ratio are widely used in population studies, they fail to capture the complexity of body composition and fat distribution, factors that are increasingly recognized as critical to cardiometabolic disease risk. Visceral fat, in particular, has been strongly associated with type 2 diabetes, cardiovascular disease, and cancer, while ectopic fat accumulation in organs such as the liver and pancreas is linked to insulin resistance, hepatic steatosis, and systemic inflammation^66,67^

To overcome the limitations of anthropometric proxies, we leveraged imaging-based assessments of fat distribution using 2D abdominal MRI and full-body DXA scans from the UKB. MRI offers high-resolution views of soft tissues and is considered the gold standard for quantifying visceral and organ-specific fat. The UK Biobank liver and pancreas imaging protocols include single-slice MRIs and shMOLLI-derived sequences that enable precise measurements of liver and pancreatic structure and composition. These images originally in DICOM format were preprocessed to 200×200 pixel JPEGs per individual for liver (FID: 20204, n = 81,496) and pancreas (FID: 20259, n = 71,169) MRIs.

For body composition analysis, we utilized whole-body DXA scans (FID: 20158, n = 83,426), which provide comprehensive measures of fat, lean mass, and bone density. We selected full-body opaque DICOM images and cropped them to focus on the upper body region (torso), producing standardized 300×300 pixel grayscale JPEGs per participant.

As shown in **Supplementary Figure 3a**, we employed a separate 2D ResNet-50 architecture for each modality liver MRI, pancreas MRI, and DXA using transfer learning to adapt the network for CAD classification. All models were trained with inputs resized to 224×224 and converted to 3-channel images to match ResNet-50 input specifications^68^. Initially, we fine-tuned models pretrained on ImageNet, a general-purpose computer vision dataset^48^. However, due to the limited domain relevance of ImageNet to medical imaging, we also tested models pretrained on RadImageNet, a large-scale medical imaging dataset comprising over 1.3 million annotated CT, MRI, and ultrasound scans across a variety of anatomical regions^47^.

As illustrated in **Supplementary Figure 3b**, RadImageNet pretraining led to marked improvements in CAD classification performance, particularly for liver and pancreas MRI, where both accuracy and F1 scores improved substantially compared to ImageNet. For liver MRI, RadImageNet-pretrained models achieved an accuracy and F1 of 0.65, compared to 0.50 and 0.42, respectively, with ImageNet pretraining. Pancreas MRI models saw similar gains (Accuracy/F1: 0.64/0.64 vs. 0.53/0.50), and DXA also benefited modestly from domain-specific pretraining (see **Supplementary Table 3**).

Model training was performed using the FastAI framework. Each modality-specific model was independently fine-tuned over 50 epochs with augmentation strategies (random flips, rotations, brightness shifts) and early stopping based on validation loss via the SaveModelCallback() method. This ensured the network adapted optimally to modality-specific features while mitigating overfitting. Together, these findings underscore the predictive utility of 2D imaging biomarkers particularly liver and pancreas fat distribution and the advantage of medical domain pretraining for CAD risk prediction.

### Embedding Extraction, Dimensionality Reduction, and Benchmarking

#### Imaging Embedding Extraction

To leverage the structural and functional richness of medical imaging, we extracted latent feature embeddings from fine-tuned deep learning models, trained separately for each imaging modality. For 3D modalities including long-axis cardiac MRI (2-, 3-, and 4-chamber views), aortic distensibility MRI, and blood flow imaging, we employed a 3D ResNet-18 model pretrained on volumetric medical datasets via MedicalNet^42,43^. Feature vectors were extracted from the final global pooling layer, yielding 512-dimensional embeddings per image. These embeddings capture dynamics such as myocardial contraction, chamber volume changes, and aortic elasticity.

For 2D imaging modalities including liver and pancreas T1 shMOLLI MRI, and whole-body DXA scans, we fine-tuned a 2D ResNet-50 model pretrained on the RadImageNet medical imaging dataset^45,47^. Each image yielded a 2048-dimensional feature embedding from the final pooling layer, capturing tissue-specific attributes such as hepatic steatosis, pancreatic fat infiltration, and body composition. Embedding extraction was performed per scan, and all features were stored for downstream analysis and multimodal integration.

#### Dimensionality Reduction via Principal Component Analysis (PCA)

To mitigate overfitting and reduce computational complexity in downstream modeling, we applied Principal Component Analysis (PCA)^41^ independently to embeddings from each imaging modality. PCA was performed on the training dataset only, and the learned transformation was applied consistently to validation and test sets.

We evaluated cumulative explained variance and discriminative performance across varying numbers of retained components (5, 10, 15, 20, and 25 PCs). Across modalities, the top 10 principal components captured the majority of informative variance (e.g., 60.79% for PC1 and 13.14% for PC2 in 4-chamber cardiac MRI; 81.19% for PC1 in aortic distensibility MRI). We selected the top 10 PCs per modality (80 total across 8 modalities) for final modeling based on the performance–dimensionality trade-off.

Critically, we observed clear separation of CAD cases from controls along PC1 in most cardiac modalities, suggesting that these compressed features retained disease-relevant structure while reducing susceptibility to overfitting as shown in **Supplementary Fig. 4**.

#### Comparison with Conventional Imaging-Derived Phenotypes (IDPs)

To benchmark the PC-based embeddings, we assembled a set of conventional imaging-derived phenotypes (IDPs) widely used in cardiovascular research and clinical practice. IDPs were extracted from UK Biobank long-axis cardiac MRI and aortic distensibility MRI using established segmentation and biomechanical modeling pipelines^69^. These included:

- **Volumetric/functional metrics**: Left ventricular end-diastolic and end-systolic volumes (FID: 24101, 24100), ejection fraction (FID: 24103), stroke volume (FID: 24102), and myocardial mass (FID: 24105).
- **Strain-based features**: Global longitudinal, circumferential, and radial strain (FID: 24174, 24157, 24181).
- **Wall thickness**: Mean myocardial wall thickness (FID: 24140).
- **Vascular stiffness**: Ascending and descending aortic distensibility (FID: 24120, 24123), derived from area changes over the cardiac cycle.

All IDPs were standardized prior to modeling. These hand-engineered metrics represent the current gold standard for structural and functional cardiac evaluation and served as a rigorous benchmark for comparison.

#### Modeling Framework for Benchmarking

We trained logistic regression classifiers under three configurations:

- **IDP-only** model using the curated set of expert-defined biomarkers,
- **PC-based** model using 10 PCA-reduced embeddings from long axis and aortic distensibility MRI (40 total),
- **Combined** model using both IDPs and PC-based features.

Model performance was evaluated using area under the receiver operating characteristic curve (AUC), and statistical comparisons between models were performed using likelihood ratio tests.

#### Canonical Correlation Analysis for Embedding Interpretability

To assess whether the deep learning models encode physiologically relevant information, we applied canonical correlation analysis (CCA)^52^ to compare the raw embeddings with conventional imaging-derived phenotypes (IDPs). CCA finds paired linear projections of two multivariate datasets that are maximally correlated, allowing us to quantify shared structure between latent embeddings and expert-defined cardiac traits.

This analysis was performed separately for each imaging modality: 2-chamber (2CH), 3-chamber (3CH), and 4-chamber (4CH) long-axis cardiac MRI, as well as aortic distensibility MRI. For each modality, we computed CCA between the 512-dimensional raw embeddings and the corresponding IDP set. The IDPs included ejection fraction, ventricular volumes, myocardial mass, strain metrics, wall thickness, and aortic distensibility traits where applicable. All features were standardized prior to analysis.

We focused on the first two canonical variable pairs (CV1 and CV2), which capture the dominant axes of shared variance. Across all four views, CV1 exhibited strong correlations between embedding and IDP projections (r = 0.816 for aortic distensibility, 0.812 for 2CH, 0.789 for 3CH, and 0.841 for 4CH), indicating that the embeddings capture a substantial amount of phenotypic information. CV2 correlations were moderate (r ≈ 0.56–0.60), suggesting that secondary axes of shared variance exist but are less dominant. These relationships are visualized in **Supplementary Figure 5**, where dot plots show the correspondence between embedding and IDP projections for each canonical variable.

To further interpret CV1, we extracted the canonical loadings of each IDP, which represent their weight on the shared axis. Loadings were visualized as bar plots for each view. This revealed that the deep models consistently prioritize contractile and volumetric features such as ejection fraction, stroke volume, and myocardial mass, with some variation by view. For example, 4CH embeddings placed higher weight on wall thickness and strain traits, while the aortic model surprisingly emphasized cardiac features more than vascular ones. These loadings provide a biologic interpretation of what the embedding space represents and how different types of physiologic signals are captured across MRI planes.

Together, these analyses indicate that the CAD-trained deep learning models are not solely encoding labels but are also learning latent representations that reflect general cardiac structure and function. CCA provides a quantitative link between deep features and conventional phenotypes, enabling interpretable comparisons and highlighting which biologic domains are most prominent in the learned representations.

### GWAS and PRS Construction and Evaluation

#### Quality Control and Genetic Data Processing

Genetic analyses were restricted to White British individuals (FID: 22006) identified using ancestry principal components (FID: 21000). Participants were excluded if they exhibited discrepancies between reported and genetic sex (FID: 22001), evidence of sex chromosome aneuploidy (FID: 22019), outlier heterozygosity or missingness (FID: 22027), genotype missingness greater than 2% (FID: 22005), or excessive relatedness defined as more than nine third-degree relatives (FID: 22021). After quality control, 7,283,324 SNPs remained. For REGENIE step 1, we applied the same filters to non-imputed genotype calls (FID: 22418) and performed LD pruning (--indep-pairwise 100kb 0.8), yielding 468,845 variants.

#### GWAS and Meta Analysis

A GWAS for coronary artery disease was then performed in 350,658 non-imaged UKB participants using REGENIE^70^ v2.2.4. Step 1 constructed genetic predictors from LD-pruned genotypes, and step 2 tested associations between imputed SNP dosages and CAD status.

Covariates included age, sex, age squared, sex–age interaction, and the first 20 UKB genetic principal components (FID: 22009). Individuals in the imaging cohort (n = 58,780) were excluded from the GWAS to prevent overlap. This GWAS included 42,115 CAD cases and 308,542 controls.

To increase power, we incorporated external GWAS summary statistics. CAD GWAS data were obtained from FinnGen^50^, which included 63,307 CAD cases and 416,171 controls, the Million Veteran Program (MVP), which provided both a European ancestry CAD GWAS^51^ with 124,302 cases and 300,039 controls and a multi-ancestry meta-analysis, and the Biobank of Japan CAD GWAS^71^, which included 29,319 cases and 183,134 controls. Two meta-analyses were then conducted using METAL. The first was a European meta-analysis that combined the non-imaged UK Biobank CAD GWAS, the FinnGen CAD GWAS, and the MVP European CAD GWAS. The second was a multi-ancestry meta-analysis that integrated the non-imaged UK Biobank CAD GWAS, the FinnGen CAD GWAS, the MVP multi-ancestry CAD GWAS, and the Biobank of Japan CAD GWAS.

#### PRS Construction and Evaluation

Polygenic risk scores were generated for the UKB imaging cohort using PRS-CS^49^ with HapMap3^72^ variants and continuous shrinkage priors. We constructed four scores. The first was a UKB-specific PRS derived from the non-imaged UKB GWAS. The second was a European meta-analysis PRS based on the combined UKB, FinnGen, and MVP European GWAS. The third was a multi-ancestry PRS using the broader meta-analysis that also included the Biobank of Japan. The fourth was GPS_Mult_^13^, a previously published multi-ancestry and multi-trait score for CAD.

Predictive performance was evaluated using Cox and Snell R² on the observed scale and liability-scale R². As shown in Figure 5, the European meta-analysis PRS achieved the highest performance (Cox and Snell R² = 0.0217; Liability R² = 0.00104), followed closely by the multi-ancestry PRS (0.0206 and 0.00099) and GPS_Mult_ (0.0194 and 0.00093). The UKB-specific PRS performed substantially worse (0.0109 and 0.00052). Based on these results, the EUR meta PRS was selected for downstream multimodal modeling.

### Electrocardiogram data

The next modalities we incorporated were the heart’s functional features, captured through Electrocardiogram (ECG). From the ECG, we utilized machine-captured metrics, including PQ Interval, P Duration, QRS Duration, QT Interval, QTC Interval, RR Interval, PP Interval, Mean P Amplitude, Mean Q Amplitude, Mean R Amplitude, and Mean T Amplitude. These measurements comprehensively capture the functional aspects of the heart.

### Metabolic and Demographic Data

We incorporated multiple metabolic measures that are well-established biomarkers for CAD risk, including Low-Density Lipoprotein (LDL), High-Density Lipoprotein (HDL), triglycerides, total cholesterol, apolipoprotein A (apoA), and apolipoprotein B (apoB). These metabolic markers are critical because they reflect lipid and protein levels associated with cardiovascular health, directly influencing plaque buildup and arterial health. We excluded lipoprotein(a) [Lp(a)] due to incomplete data for many individuals.

Additionally, we included demographic traits known to correlate with CAD risk: age, sex, Body Mass Index (BMI), average household income, Townsend deprivation index (a measure of socioeconomic status), Diabetes Mellitus status, and smoking status. These demographic factors are essential as they provide context for lifestyle and socioeconomic influences on heart health, enhancing the model’s predictive power for CAD. We also added self-reported medication history for insulin, lipid lowering and blood pressure lower drugs.

### Multimodal Risk Modeling and Feature Selection

To construct a robust predictive model for CAD, we integrated diverse biomedical data types into a unified multimodal dataset. The feature set included imaging-derived principal components (PCs), PRS, metabolic biomarkers, ECG-based functional metrics, and baseline demographic and clinical information. The imaging features were obtained from PCA-transformed deep learning embeddings of eight distinct modalities: 2-, 3-, and 4-chamber long-axis cardiac MRI, aortic distensibility MRI, blood flow MRI, liver MRI, pancreas MRI, and DXA scans. Each modality contributed 10 principal components, resulting in 80 imaging features. Demographic and clinical variables included age, sex, body mass index (BMI), smoking status, blood pressure, medication history and diabetes status. Metabolic biomarkers comprised serum measurements of LDL, HDL, total cholesterol, triglycerides, apolipoprotein A (apoA), and apolipoprotein B (apoB). Genetic predisposition was quantified using a PRS trained on GWAS from three different European cohorts. Socioeconomic features included household income and Townsend deprivation index. ECG-derived features such as PR, QRS, QT, and RR intervals and waveform amplitudes were also included to capture cardiac conduction and rhythm phenotypes. Imaging center was encoded as a covariate to adjust for site-specific biases.

Participants with missing data for any feature group were excluded from the analysis to maintain consistency and avoid reliance on imputation. Most exclusions stemmed from unavailable PRS (due to non-European ancestry and participant removal during genotyping QC) and missing lipid profiles. After filtering, the final training dataset consisted of 3,000 CAD cases and 3,000 matched controls. An independent, balanced test set of 200 incident CAD cases and 200 matched controls, not used during training or hyperparameter tuning, was held out for final evaluation. In addition, to evaluate model performance under realistic prevalence conditions, we assessed all classifiers on a separate imbalanced test set comprising 200 incident CAD cases and 3,000 controls.

To quantify the individual predictive capacity of each feature group, we trained L1-regularized logistic regression (LASSO) models using only one group at a time. All models were evaluated using five-fold cross-validation, stratified to maintain class balance in each fold. Model performance was quantified using Nagelkerke’s pseudo R_N_², which estimates the proportion of variance in the outcome (CAD status) explained by the model. To provide robust estimates of variability, we applied bootstrap resampling with 1,000 iterations, computing 95% confidence intervals for each R_N_² value. These analyses allowed us to rank feature groups by their standalone predictive utility.

To evaluate how each group contributed to overall predictive performance in a multimodal setting, we trained a full logistic regression model using all available features. We then implemented a leave-one-out (LOO) feature ablation approach: each feature group was excluded from the full model one at a time, and model performance was recomputed. The difference in R_N_² between the full and reduced models was used to estimate the marginal contribution of the omitted group. As with the standalone models, bootstrapped confidence intervals were calculated to assess the robustness of these estimates.

To further investigate the degree of redundancy and complementarity among feature groups, we performed a commonality analysis. This technique decomposes the explained variance of a full model into unique and shared components for each feature group. First, we trained models using each group independently to estimate total variance explained. We then incrementally introduced additional one at a time groups (ECG-derived features, metabolic biomarkers, and baseline demographic traits) and recomputed the R_N_² at each step. The change in explained variance was used to determine the proportion of unique signal captured by the added group versus variance shared with previously included features. This approach enabled us to distinguish groups that contributed novel information from those whose signal overlapped with common phenotypes like BMI.

To test model architectures for CAD classification, we evaluated a suite of supervised learning methods including logistic regression, LASSO logistic regression, support vector machines, gradient boosting machines (GBM), XGBoost, LightGBM, and random forests. Each model was trained on the full multimodal dataset using five-fold stratified cross-validation. All continuous features were z-score normalized, and categorical variables were one-hot encoded. Model performance was evaluated using both cross-validation and independent test set performance, assessed via accuracy and area under the receiver operating characteristic curve (AUC). Results of model benchmarking are reported in **Supplementary Fig. 7.** The final model used for downstream classification was selected based on the best trade-off between predictive performance and interpretability, with preference given to models that supported transparent feature attribution.

### Multimodal Survival Modeling and Imaging-Enhanced Risk Stratification

#### Cohort Design and Time-to-Event Definition

To assess the prognostic value of multimodal features for long-term CAD risk, we conducted a time-to-event analysis using longitudinal data from the UK Biobank (UKB). The outcome of interest was the incidence of CAD within 10 years following the baseline imaging assessment. Time-at-risk was defined as the interval between the imaging visit and the earliest recorded diagnosis of CAD, as captured through hospital inpatient records and death registry linkage.

Participants who remained event-free during follow-up were administratively censored at 10 years. Individuals with a known CAD diagnosis prior to their imaging visit were included in the model but treated as prevalent cases, with their time-at-risk set to a small positive value (0.001 years) to enable modeling while minimizing their impact on time-to-event estimation. These individuals were excluded from survival curve visualizations and stratified risk group analyses, which focused exclusively on incident outcomes.

The modeling cohort combined participants from both the training and held-out test datasets described previously, yielding a total sample of 6,400 individuals after excluding those with missing feature values or non-European ancestry (to align with PRS applicability). This ensured consistency across modeling pipelines and maximized statistical power for evaluating hazard-based outcomes.

#### Feature Set Construction and Preprocessing

We constructed four feature groups representing distinct domains of CAD risk:

1. **Baseline clinical features**: age, sex, body mass index (BMI), and smoking status.
2. **Metabolic biomarkers**: total cholesterol, low-density lipoprotein (LDL), triglycerides, apoA, apoB, medication history and a binary indicator for diabetes mellitus status.
3. **Genetic risk**: PRS derived from a multi cohort European meta GWAS-based model capturing inherited predisposition to CAD.
4. **Imaging-derived features**: 40 principal components extracted from deep learning embeddings of long-axis cardiac MRI (2-, 3-, and 4-chamber views) and aortic distensibility MRI (10 PCs per modality).

All continuous variables were z-score normalized prior to modeling to ensure comparability across scales. Categorical features were one-hot encoded as appropriate. Imaging embeddings were derived using fine-tuned convolutional neural networks as previously described, with PCA applied separately per modality to retain the most variance-explaining components. The resulting compressed features were used in all survival analyses to represent subclinical structural and functional information.

#### Cox Proportional Hazards Modeling

To evaluate the incremental contribution of genetic and imaging data, we trained three nested Cox proportional hazards models using five-fold cross-validation. These models were:

- **Clinical biomarker model**: incorporated baseline clinical features, metabolic biomarkers, and diabetes status, aligned with the traditional Framingham Risk Score structure^3^.
- **Clinical biomarker + PRS model**: included all features from the Framingham model plus PRS.
- **Full multimodal model**: added imaging-derived PCs to the Framingham + PRS model.

Each model was evaluated using the concordance index (C-index), which quantifies the probability that the model correctly ranks the survival times of any two randomly chosen individuals. Cross-validation folds were stratified to preserve event proportion, and model performance was averaged across folds. Improvement in the C-index across nested models was used to quantify the marginal value of each data modality for long-term CAD risk prediction.

#### Hierarchical Risk Stratification Framework

To examine the interpretability and clinical applicability of our multimodal model, we implemented a three-step hierarchical stratification strategy. This framework mimics how genetic and imaging data might be deployed sequentially in real-world clinical workflows:

1. **Clinical stratification**: Participants were first stratified into tertiles (low, medium, and high risk) based on hazard scores generated from the clinical-only model. These tertiles approximate clinical categories used in standard cardiovascular screening.
2. **Genetic stratification**: Among individuals in the highest clinical tertile, a second stratification was applied using PRS values. These participants were again divided into low, medium, and high genetic risk groups based on PRS tertiles.
3. **Imaging-based stratification**: Within the high clinical + high PRS subgroup, we trained a separate Cox model using only imaging-derived PCs to generate an imaging-specific risk score. This score was used to assign individuals into low, medium, and high imaging risk tertiles.

Survival curves for each tier of stratification were estimated using Kaplan–Meier methods, and statistical differences between curves were assessed using log-rank tests. This approach enabled us to evaluate how well each layer of information clinical, genetic, and imaging refined risk predictions for individuals already considered at high risk under conventional metrics.

#### Relative Risk Estimation

To quantify the degree of stratification afforded by each risk layer, we calculated relative risk (RR) for each risk group at every tier. RR was defined as the ratio of the incidence rate of CAD in a given group to that in the low-risk group identified by the clinical-only model. This was repeated after each stratification step to assess how genetic and imaging features sharpened risk differentiation. The topmost group (high clinical+ high PRS + high imaging risk) exhibited more than a fifteen-fold increase in event incidence relative to the baseline, underscoring the power of multimodal risk modeling to surface individuals with extreme vulnerability who would otherwise be overlooked.

### Spatial Cross-Validation to Evaluate Site-Level Generalizability

To evaluate whether our CAD prediction framework generalized across independently acquired datasets, we employed a spatial cross-validation design using the UK Biobank’s multi-center imaging structure^53^. Imaging data were collected at four centers Bristol, Cheadle, Newcastle, and Reading each with distinct scanner systems, technician teams, and participant subpopulations.

These site-level variations provided a natural test bed for generalization without relying on an external dataset.

We applied a leave-one-center-out cross-validation scheme. For each of four folds, we withheld one center’s data for testing and trained our full multimodal modeling pipeline on the remaining three. This approach was applied independently for both binary classification and survival prediction pipelines.

#### Multimodal Classification Pipeline

For classification, we began by fine-tuning 3D ResNet-18 models (pretrained via the Med3D framework) on long-axis cardiac MRI and aortic distensibility MRI data from the training centers. For each participant, we extracted imaging embeddings from the final global pooling layers of these models and applied Principal Component Analysis (PCA) to retain the top 10 components per modality. These imaging-derived PCs were concatenated with baseline demographic data (age, sex, BMI), metabolic biomarkers (LDL, HDL, triglycerides, total cholesterol), smoking status, and diabetes status to form a unified feature vector.

An XGBoost classifier was trained using this multimodal feature set. The model was evaluated on the held-out center’s data, and predictive performance was measured using the area under the receiver operating characteristic curve (AUC). All numerical features were z-score normalized prior to training. Feature distributions were visualized across centers to confirm consistency in data preprocessing.

#### Survival Modeling Pipeline

To mirror the classification setup, we implemented spatial cross-validation for survival analysis using Cox proportional hazards models. The same leave-one-center-out design was applied. The multimodal feature set was identical to that used for classification, with time-to-event outcomes defined as the duration from imaging visit to incident CAD diagnosis or censoring at 10 years.

For each fold, a Cox model was trained on participants from three centers and tested on the fourth. Performance was quantified using the concordance index (C-index) to assess how well the model ranked individuals by risk of CAD events. Consistency in C-index across centers indicated that the survival model maintained reliability even when exposed to site-level heterogeneity.

#### End-to-End Pipeline Validation

This spatial cross-validation strategy tested every stage of our modeling pipeline: image preprocessing, feature extraction, dimensionality reduction, multimodal fusion, and outcome prediction. The observed stability in both AUC and C-index across geographically distinct centers underscores the robustness of our approach and its suitability for deployment across diverse clinical settings. These results lay a strong foundation for future validation studies using fully external datasets.

## Computing Infrastructure

All computational analyses were performed on the Corral and Lonestar6 systems at the Texas Advanced Computing Center (TACC). Deep learning experiments were conducted using three NVIDIA A100 GPUs with 40GB HBM2 memory, utilizing CUDA toolkit version 11.1.

## Data Availability

All data are made available from the UKB (https://www.ukbiobank.ac.uk/enable-your-research/apply-for-access) to researchers from universities and other institutions with genuine research inquiries following institutional review board and biobank approval. This research has been conducted using the UKB resource under application number 65439.

## Code Availability

The code used for analysis and figure generation is available via GitHub at https://github.com/Devanshpandey/preCog-Multimodal-AI-for-Precision-Cardiology

## Acknowledgements

Computing support for the project was provided through the Director’s Discretionary Fund at the Texas Advanced Computing Center (TACC). V.M.N. was supported by a grant for human brain evolution through the Allen Discovery Center program, a Paul G. Allen Frontiers Group advised program of the Paul G. Allen Family Foundation, as well as by a Good Systems Fellowship for Ethical AI at The University of Texas at Austin.

## Contributions

D.P. and V.M.N. conceived and designed the study. Methodological development and analytical strategies were jointly led by D.P. and V.M.N. Data analysis, and investigation were carried out by D.P., L.X., J.Y.W., E.K., C.L., A.M. and V.M.N. Visualization and figure generation were performed by D.P. and V.M.N. V.M.N. secured funding and, along with D.P., coordinated overall project administration. Supervision was provided by J.C.D., E.C., J.N., C.A.T. and V.M.N., D.P. and V.M.N. co-wrote the initial manuscript draft.

## Ethics declarations

C.A.T founded HeartFlow, Inc., and currently serves as a member of the board of directors for both HeartFlow, Inc., and Ebenbuild, GmbH. Rest of the authors declare no competing interests

---

**Supplementary Figure 1:**
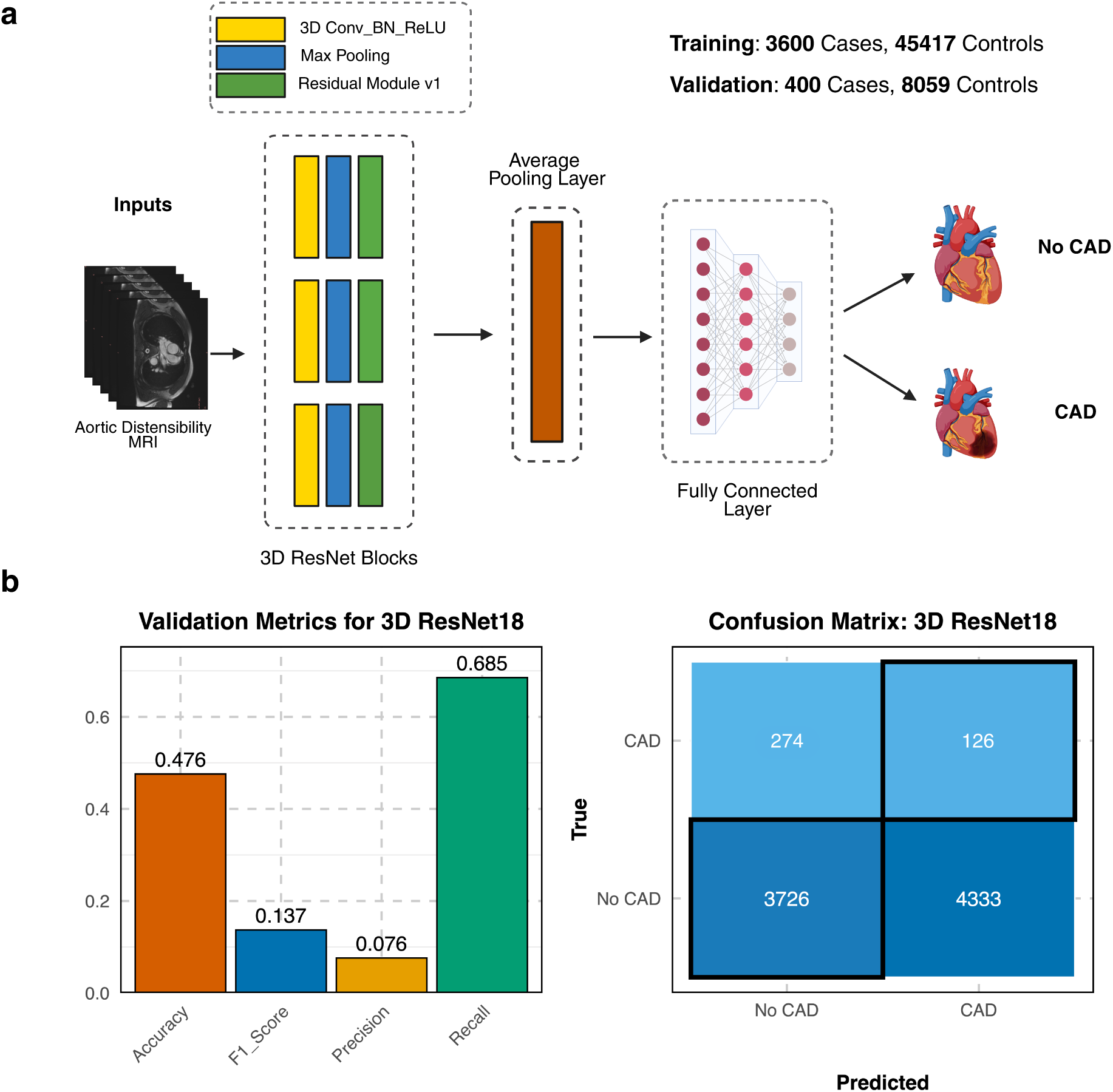
Evaluation of 3D ResNet-18 model trained on an imbalanced dataset for CAD classification. **a**, Schematic overview of the 3D ResNet-18^43^ architecture used to classify CAD status from aortic distensibility MRI volumes. The model was initialized with Kinetics-400 pretrained weights and fine-tuned using 4,000 CAD cases and 53,476 controls. **b**, Validation metrics and confusion matrix from the imbalanced model. Although the model achieved high recall (0.685), it exhibited poor precision (0.076) and F1 score (0.137), with most samples classified as CAD-positive regardless of ground truth. These results motivated the decision to retrain using a balanced dataset to improve discriminative performance and reduce prediction bias.

**Supplementary Figure 2:**
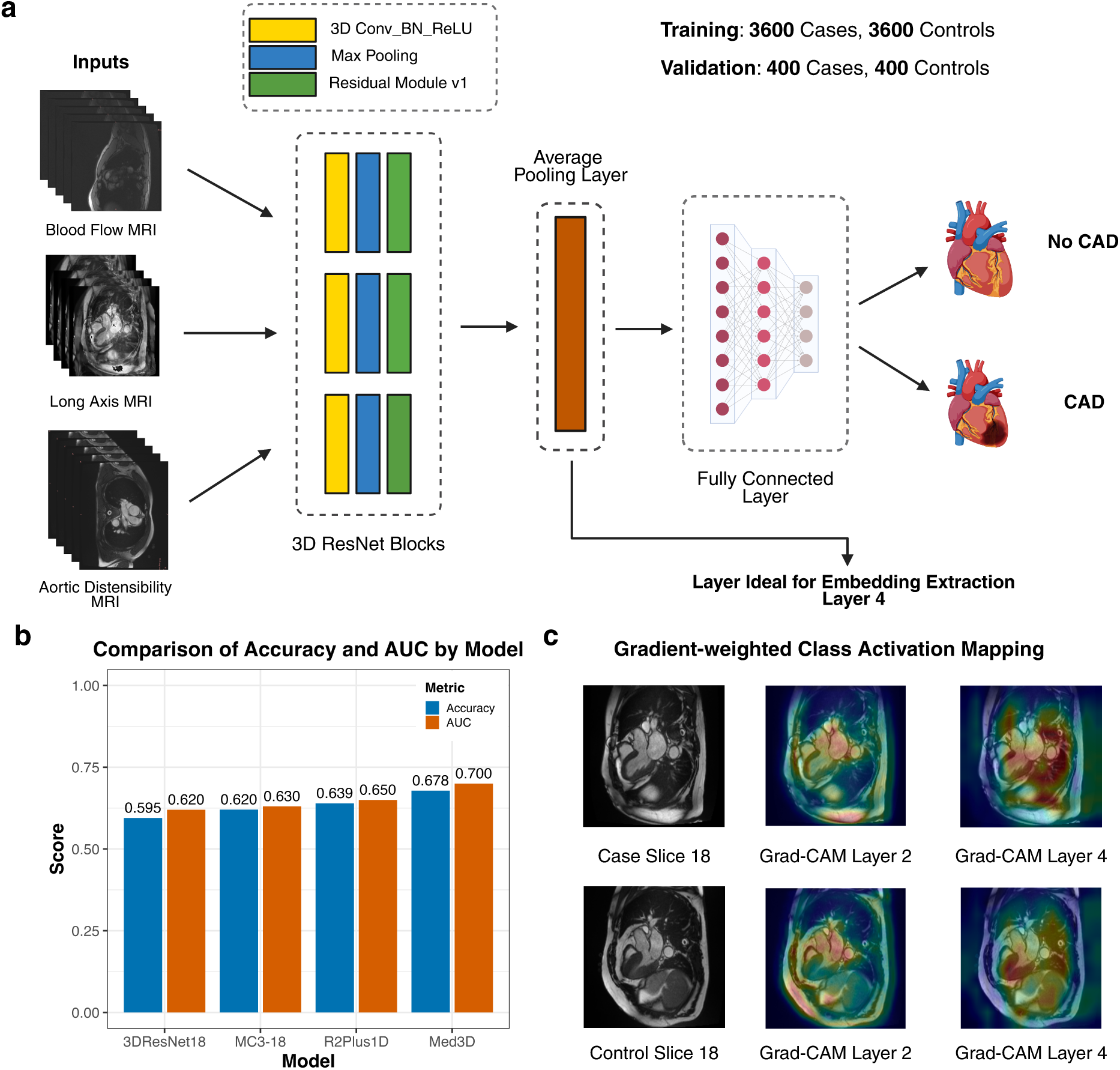
Framework for fine-tuning 3D convolutional models for CAD prediction using cardiac MRI. **a**, Schematic overview of the 3D ResNet architecture used to classify CAD from three cardiac MRI modalities: blood flow MRI, long-axis heart MRI, and aortic distensibility MRI. Independent models were trained for each modality using a 3D ResNet initialized with MedicalNet (Med3D) or Kinetics-400 pretrained weights. Training was conducted on a balanced dataset of 3,600 CAD cases and 3,600 controls, with 400 cases and 400 controls reserved for validation. The final average pooling layer was used for feature extraction, and predictions were generated via a fully connected output head. **b**, Comparison of classification performance across different 3D convolutional architectures. Med3D outperformed other pretrained models (3D ResNet-18, MC3-18, R(2+1)D) in both accuracy (0.678) and AUC (0.700), highlighting the value of pretraining on medical imaging tasks for transfer learning in clinical classification. **c**, Gradient-weighted class activation mapping (Grad-CAM) visualizations for a CAD case and control, using intermediate (layer 2) and final (layer 4) convolutional blocks. Heatmaps from the CAD-positive case highlight activation in the left ventricular wall and interventricular septum, suggesting that the model is focusing on regions associated with structural remodeling relevant to CAD pathophysiology.

**Supplementary Figure 3:**
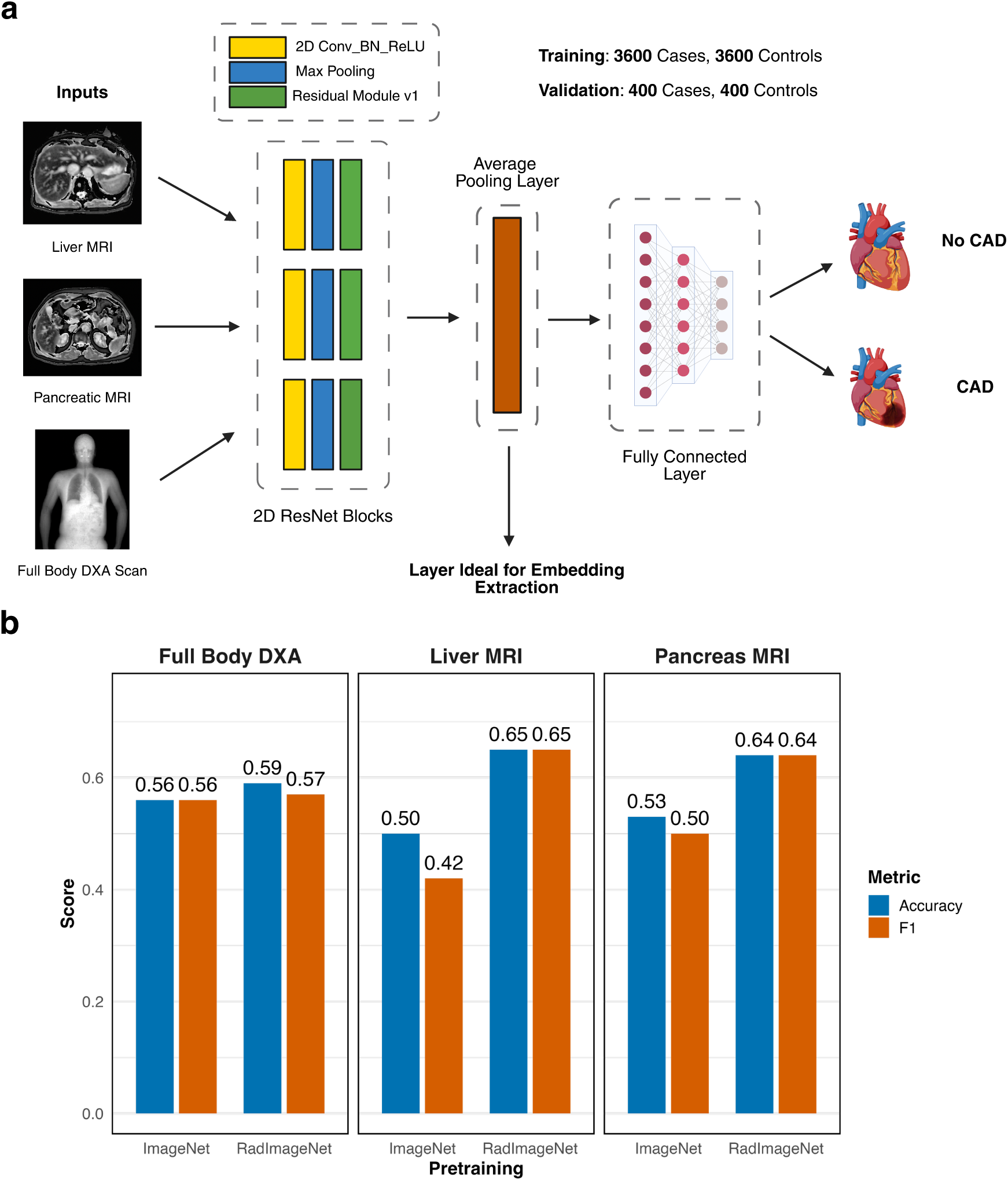
CAD Prediction Using 2D MRI and DXA Imaging with Domain-Specific Pretraining. **a**, Schematic overview of the model architecture for each 2D imaging modality. Liver MRI, pancreas MRI, and full-body DXA images were each processed through independent 2D convolutional models based on a ResNet-50 backbone. Models were initialized with either ImageNet or RadImageNet pretraining and fine-tuned for binary classification of CAD status. The final prediction was generated via a fully connected layer followed by a sigmoid activation. **b**, Comparison of classification performance across modalities and pretraining strategies. Models pretrained on RadImageNet consistently outperformed ImageNet-pretrained models, particularly for liver MRI (accuracy/F1: 0.65/0.65) and pancreas MRI (accuracy/F1: 0.64/0.64). Full-body DXA models also showed modest improvement with RadImageNet (accuracy/F1: 0.59/0.57 vs. 0.56/0.56). These results highlight the value of domain-specific pretraining for enhancing CAD prediction from 2D imaging biomarkers.

**Supplementary Figure 4:**
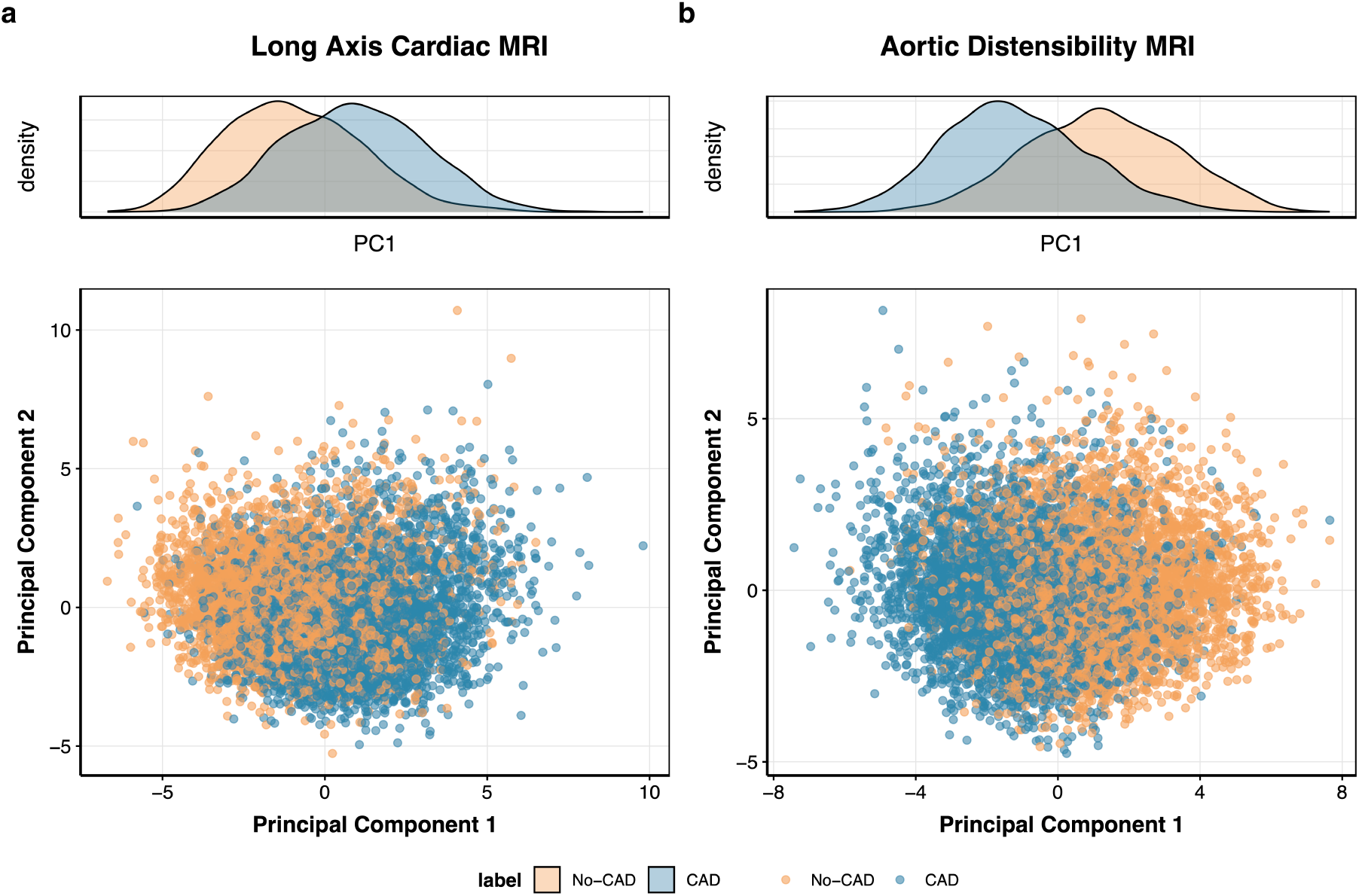
Separation of CAD cases and controls in principal component space for two cardiac imaging modalities. **a**, Long-axis cardiac MRI and **b**, aortic distensibility MRI embeddings are visualized along their first two principal components (PC1 and PC2), with CAD cases shown in blue and controls in orange. Density plots above each scatter plot show clear distributional shifts along PC1, reflecting underlying disease-related structure.

**Supplementary Figure 5:**
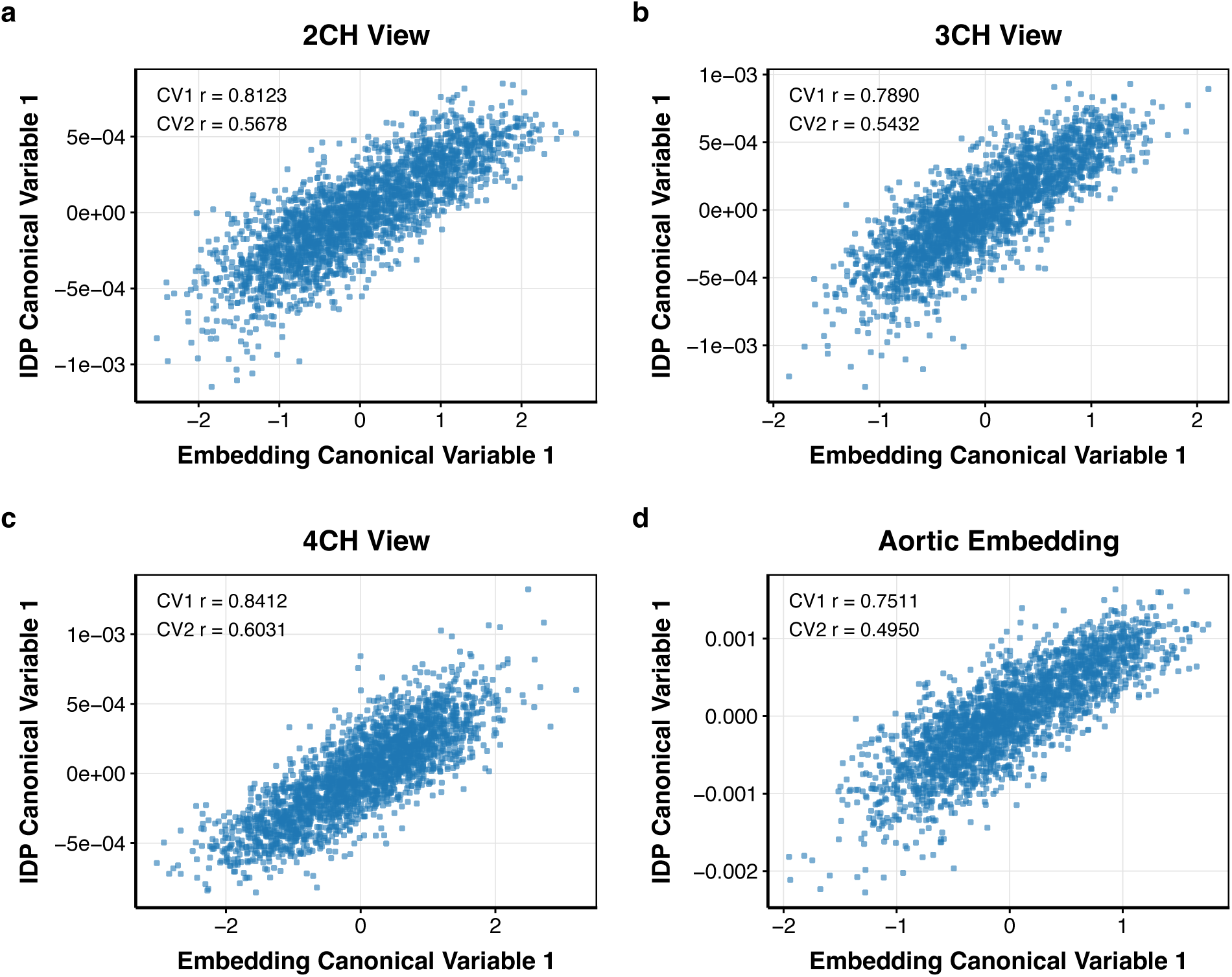
Canonical correlation analysis (CCA) reveals strong alignment between raw deep learning embeddings and conventional imaging-derived phenotypes (IDPs). Scatter plots display the first canonical variable (CV1) projections from CCA for raw embeddings (x-axis) and IDPs (y-axis) across four imaging views: **a,** 2-chamber long axis MRI (2CH), **b**, 3-chamber long axis MRI (3CH), **c** 4-chamber long axis MRI (4CH), and **d,** aortic distensibility MRI. Canonical correlation coefficients (r) for the first two canonical pairs are reported in each panel. Across all views, CV1 showed high correlation (r = 0.751–0.841), indicating that a large portion of the variance in expert-defined cardiac traits is captured by the latent representations learned by the vision models. These findings demonstrate that CAD-trained embeddings reflect biologically meaningful structure even prior to dimensionality reduction.

**Supplementary Figure 6:**
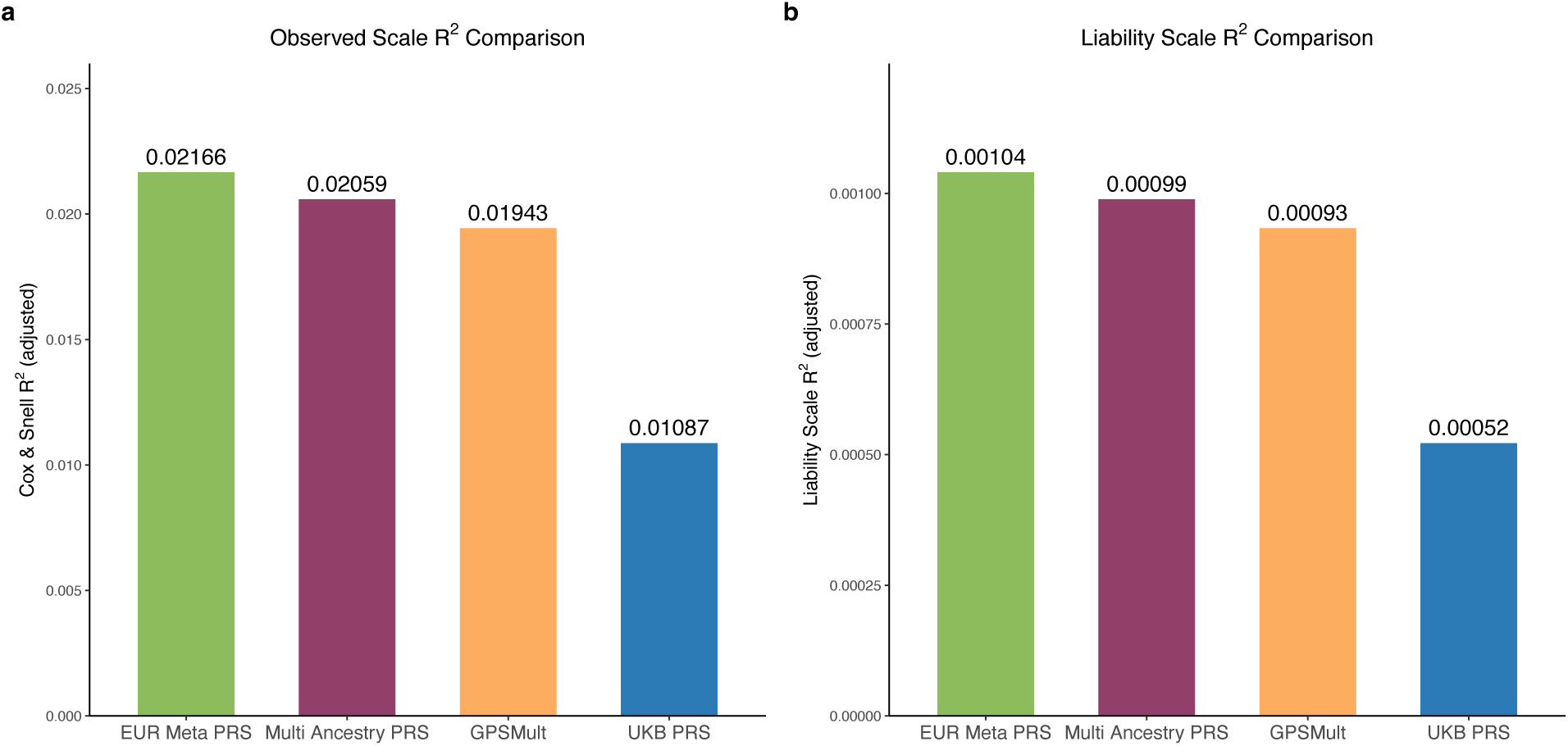
Performance comparison of polygenic risk scores for CAD. **a**, Observed-scale predictive performance, measured using Cox and Snell R² (adjusted), for four polygenic risk scores: the European meta-analysis PRS, the multi-ancestry meta-analysis PRS, GPS_Mult_, and the UKB-specific PRS. The European meta-analysis PRS achieved the highest explanatory power (R² = 0.0217), followed by the multi-ancestry PRS (0.0206) and GPS_Mult_ (0.0194), while the UKB PRS performed substantially worse (0.0109). **b**, Liability-scale R² (adjusted) comparison of the same scores. The European meta-analysis PRS again showed the strongest performance (R² = 0.00104), with similar values for the multi-ancestry PRS (0.00099) and GPS_Mult_ (0.00093), and markedly lower performance for the UKB PRS (0.00052).

**Supplementary Figure 7:**
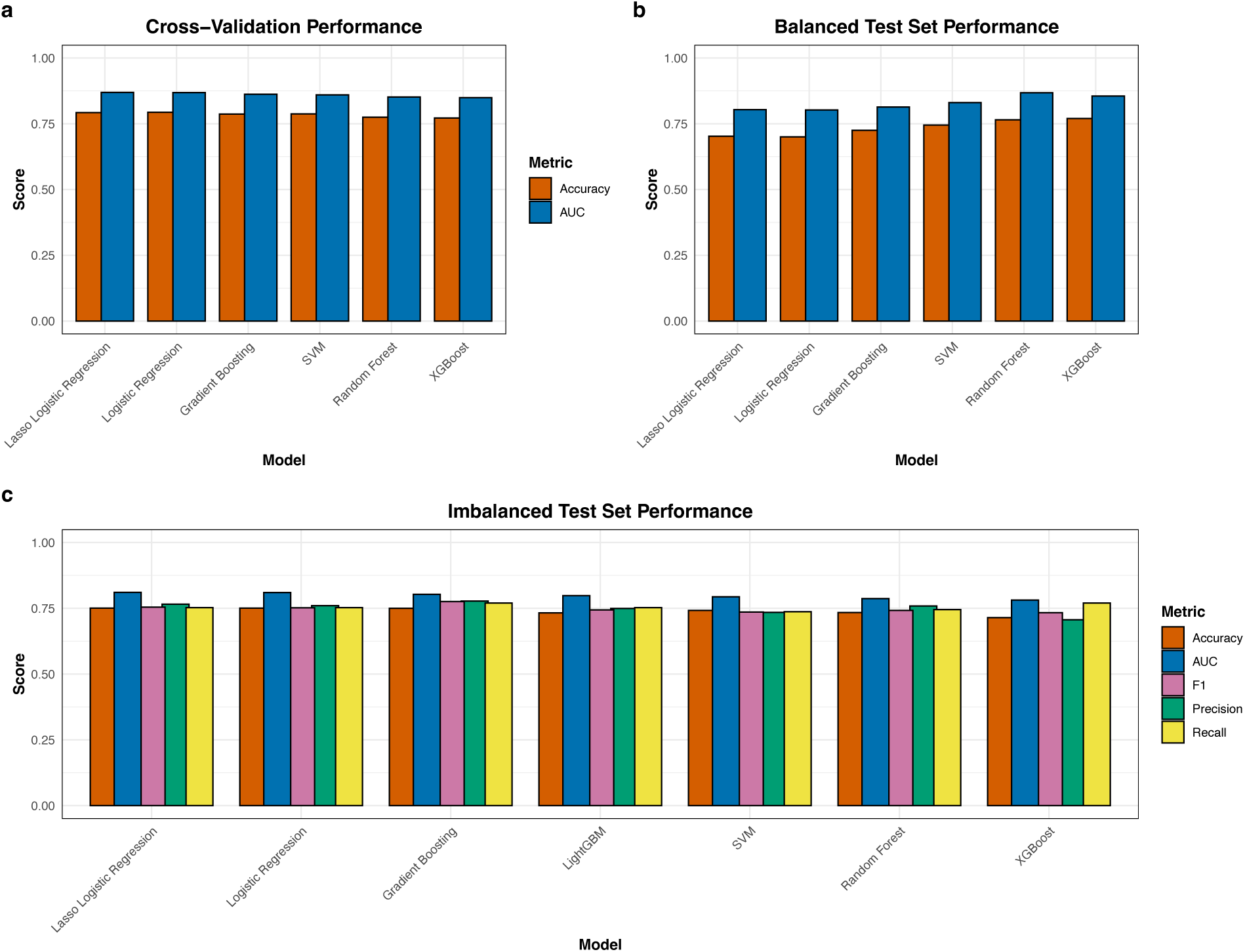
Performance comparison of supervised learning models for multimodal CAD classification. **a**, Five-fold cross-validation accuracy and AUC for each classifier trained on the integrated multimodal dataset, including PCA-reduced imaging features (from long-axis cardiac and aortic distensibility MRI), PRS, metabolic biomarkers, ECG-derived features, and demographic data. **b**, Accuracy and AUC on a held-out, balanced test set (200 incident CAD cases and 200 matched controls) for the same classifiers. **c**, Evaluation on an imbalanced test set comprising 200 incident CAD cases and 3,000 controls, showing accuracy, precision, recall, F1 score, and AUC. Models compared include logistic regression, LASSO logistic regression, gradient boosting, support vector machines (SVM), XGBoost, and random forest.

**Supplementary Table 1:** Overview of imaging modalities used in the multimodal CAD prediction framework. Each modality contributes unique anatomical or physiological insights. Sample sizes reflect the number of CAD cases and controls with available data per modality.

| Imaging Modality | Description | Sample Size |
| --- | --- | --- |
| Cardiac Long Axis MRI | Three-chamber cine MRI capturing structural dynamics of the left and right heart | 4872 cases and 65341 controls |
| Cardiac Aortic Distensibility MRI | Time-resolved MRI measuring aortic elasticity and vessel wall motion during the cardiac cycle | 4691 cases and 58576 controls |
| Cardiac Blood Flow MRI | Phase-contrast MRI capturing intraventricular blood flow patterns and hemodynamics | 4439 cases and 66499 controls |
| Liver MRI | Single-slice abdominal MRI assessing hepatic fat infiltration and tissue composition | 4451 cases and 64273 controls |
| Pancreas MRI | Single-slice abdominal MRI capturing pancreatic morphology and ectopic fat content | 4549 cases and 57623 controls |
| Full Body DXA Opaque | Whole-body dual-energy X-ray absorptiometry image assessing fat and lean mass distribution | 4587 cases and 66005 controls |

**Supplementary Table 6:** Descriptive statistics for ECG-derived features used in the multimodal CAD prediction framework. Values are reported as mean ± standard deviation (SD). All amplitudes are measured in microvolts (μV); durations are in milliseconds (ms).

| Feature | Mean ± SD |
| --- | --- |
| QRS Duration | 88.74 ± 15.06 ms |
| QT Interval | 420.71 ± 32.13 ms |
| QTC Interval | 422.78 ± 25.27 ms |
| RR Interval | 996.69 ± 166.15 ms |
| PP Interval | 989.14 ± 187.61 ms |
| Mean Q Amplitude | 219.75 ± 219.79 μV |
| Mean R Amplitude | 642.53 ± 210.68 μV |
| Mean T Amplitude | 184.00 ± 96.55 μV |

**Supplementary Table 7:** Descriptive statistics for clinical and metabolic features used in the multimodal CAD prediction framework. Values are presented as mean ± standard deviation (SD) for continuous variables or percentage breakdowns for categorical variables.

| Feature Type | Summary Statistics (Mean ± SD or Median [IQR]) |
| --- | --- |
| Age | 65.418 ± 7.803 years |
| Sex | 51.8% female and 48.2% male |
| BMI | 26.569 ± 4.228 kg/m<sup>2</sup> |
| apoA | 1.546 ± 0.265 g/L |
| apoB | 1.03 ± 0.229 g/L |
| LDL | 3.577 ± 0.828 mmol/L |
| HDL | 1.472 ± 0.377 mmol/L |
| Total Cholesterol | 5.721 ± 1.08 mmol/L |
| Triglycerides | 0.331 ± 0.196 mmol/L |
| Diastolic BP | 81.385 ± 10.452 mmHg |
| Systolic BP | 137.165 ± 18.889 mmHg |
| Pulse Rate | 67.885 ± 10.937 bpm |

**Supplementary Table 13:** Leave-one-center-out performance of the multimodal CAD prediction framework across UK Biobank imaging centers. Each fold trains the model on three centers and evaluates on the held out fourth. Sample sizes, test AUC (classification), and concordance index (C-index; survival prediction) are reported for each fold.

| Training Centers | Test Center | Number of Training Samples | Number of Test Samples | Test AUC | Test C-Index |
| --- | --- | --- | --- | --- | --- |
| Cheadle, Bristol and New Castle | Reading | 2519 cases and 2562 controls | 608 cases and 725 controls | 0.805 | 0.762 |
| Bristol, Reading, New Castle | Cheadle | 1740 cases and 1747 controls | 1387 cases and 1540 controls | 0.822 | 0.748 |
| New Castle, Reading, Cheadle | Bristol | 2897 cases and 2988 controls | 230 cases and 290 controls | 0.785 | 0.751 |
| Reading, Cheadle, Bristol | New Castle | 2225 cases and 2564 controls | 723 cases and 903 controls | 0.795 | 0.732 |

